# A prospective real-time transfer learning approach to estimate Influenza hospitalizations with limited data

**DOI:** 10.1101/2024.07.17.24310565

**Authors:** Austin G Meyer, Fred Lu, Leonardo Clemente, Mauricio Santillana

## Abstract

Accurate, real-time forecasts of influenza hospitalizations would facilitate prospective resource allocation and public health preparedness. State-of-the-art machine learning methods are a promising approach to produce such forecasts, but they require extensive historical data to be properly trained. Unfortunately, historically observed data of influenza hospitalizations, for the 50 states in the United States, are only available since the beginning of 2020, as their collection was motivated and enabled by the COVID-19 pandemic. In addition, the data are far from perfect as they were under-reported for several months before health systems began consistently and reliably submitting their data. To address these issues, we propose a transfer learning approach to perform data augmentation. We extend the currently available two-season dataset for state-level influenza hospitalizations in the US by an additional ten seasons. Our method leverages influenza-like illness (ILI) surveillance data to infer historical estimates of influenza hospitalizations. This cross-domain data augmentation enables the implementation of advanced machine learning techniques, multi-horizon training, and an ensemble of models for forecasting using the ILI training data set, improving hospitalization forecasts. We evaluated the performance of our machine learning approaches by prospectively producing forecasts for future weeks and submitting them in real time to the Centers for Disease Control and Prevention FluSight challenges during two seasons: 2022-2023 and 2023-2024. Our methodology demonstrated good accuracy and reliability, achieving a fourth place finish (among 20 participating teams) in the 2022-23 and a second place finish (among 20 participating teams) in the 2023-24 CDC FluSight challenges. Our findings highlight the utility of data augmentation and knowledge transfer in the application of machine learning models to public health surveillance where only limited historical data is available.

**Author summary:** Influenza is a major public health concern in the United States, causing thousands of hospitalizations annually. Accurate and timely forecasts of hospitalization rates are essential for effective public health preparedness. However, limited historical data makes forecasting with state-of-the-art models challenging. To address this issue, we developed a cross-domain data augmentation method that allowed us to train advanced machine learning models using symptom-based (syndromic) surveillance data. We then created a set of models, focusing on gradient-boosted machines, and combined them into an ensemble framework. This approach successfully overcame data limitations, outperforming the majority of teams participating in the CDC FluSight project for 2022-23 and 2023-24. Additionally, our forecasts demonstrated superior accuracy to the CDC’s composite model in the 2022-23 season and matched its performance in 2023-24. Our study demonstrates a robust and data-efficient strategy for training machine learning models for use in public health forecasting.

## Introduction

Influenza is a viral respiratory infection that can cause severe illness in humans. While it has circulated in humans for several centuries, it continues to pose a significant challenge to global public health, leading to substantial annual morbidity and mortality [1]. In the United States, for example, annual influenza-related hospitalizations range from 100,000 to 700,000, resulting in 5,000-50,000 deaths [2, 3]. The availability of accurate, real-time forecasts of influenza activity across geographies would enhance preparedness and response strategies for public health agencies, medical institutions, businesses, and the general public. Yet, the unpredictable nature of influenza epidemics and the limited historical surveillance data make effective forecasting a challenge [4, 5].

The field of influenza forecasting has evolved significantly in recent years, with a marked increase in research efforts combining multiple data sources with a blend of methodologies to forecast influenza at the state level [6–8]. A wide variety of statistical methods are effective at this task, including regularized and Bayesian regression [7, 9–11], autoregressive methods [12], and non-parametric approaches [13, 14]. Mechanistic and compartmental approaches are also widely used for influenza forecasting [8, 15–17]. In addition, the increase in data availability has led to a rise of newer machine learning methods [18, 19].

The US Centers for Disease Control and Prevention (CDC) FluSight challenges have played a pivotal role in the advancement of this field since 2013 by inviting the scientific community to develop and evaluate forecasting models prospectively and in real-time [20–22]. Recently, FluSight moved away from requesting forecasts of influenza activity as captured by syndromic surveillance—i.e. systems that capture the number of patients seeking medical attention for influenza-like illnesses (ILI)—instead pivoting to forecasts of hospitalizations attributable to influenza. Given the connection to health system utilization, this metric is more relevant for public health officials, and its availability was made possible by the enhanced data collection following the start of the COVID-19 pandemic and the limitations in forecasting case counts inferred from symptoms-based disease surveillance [23]. Unfortunately, prior to 2020, influenza hospitalization data was only consistently available in about seven states through the CDC’s Emerging Infections Program (EIP).

Unfortunately, the recent shift to focusing on monitoring and forecasting hospitalizations means that there are only two influenza seasons (2020-21 and 2021-22) with ground-truth hospitalization data available from all 50 states. Furthermore, in these two years the hospitalization curves were highly atypical due to the COVID-19 pandemic, meaning that there is almost no useful data for modeling hospitalizations. This data sparsity presents a significant limitation to training forecasting models. While traditional statistical models like ARIMA and exponential smoothing can be trained with limited data, their forecasting accuracy suffers as the calibration of hyper-parameters, such as the selection of lag orders for the model components, are unstable with the limited training data. Similarly, robust multivariate statistical models like vector autoregression (VAR) struggle to converge with a large number of unknowns relative to a limited set of observations in the training set. In this context, data-intensive machine learning models like gradient-boosted machines and neural networks simply cannot be trained at all with under two seasons of training data.

Beyond the data sparsity issue, creating real-time prospective forecasts for influenza hospitalizations at a cadence useful to public health officials, presents significant challenges. For example, at the start of the 2022-23 season, we were given less than two weeks to begin forecast submission; thus, we had just two weeks to develop a solution to the data sparsity issue. Moreover, the real-time nature of the prediction occasionally requires last-minute adjustments to the scope of the challenge—for instance, this occurred during the 2023-24 season due to changes in the availability of hospitalization data. In addition, there are issues with data reporting, potential data coverage gaps, and variability in data quality across different regions that collectively make it challenging to create accurate and reliable real-time forecasts, particularly when historical data is limited.

In addressing the challenges presented by limited data availability and the need for accurate forecasts, our study introduces an innovative solution. Leveraging the transfer learning approach of applying knowledge from one domain to another, we first designed a cross-domain data augmentation strategy to utilize prior symptom-based surveillance data across all US states. Specifically, we model historical influenza hospitalizations using ILI surveillance data, creating an enriched dataset that adds a decade’s worth of estimated hospitalizations for every state or territory where ILI data are available (all except Florida and Puerto Rico). Next, we used this large transformed dataset to perform knowledge transfer at the start of the 2022-23 season, enabling us to develop and tune state-of-the-art machine learning methodologies to forecast flu hospitalization at a time when usable hospitalization data was scarce. This approach enabled us to develop accurate state-specific and national forecasting models, including ARIMA, VAR, and gradient-boosted machine learning techniques (e.g., LightGBM) [24]. These models were integrated into an ensemble framework that was rigorously, verifiably, and prospectively validated in the CDC’s FluSight challenge over two seasons (2022-23 and 2023-24). Overall, our data augmentation, machine learning, and ensembling placed us fourth in 2022-23 and second in 2023-24 among more than 30 models in a national challenge and also allowed us to be among the four teams to exceed the CDC’s combined weighted ensemble in any season.

## Materials and methods

### Data availability

#### Influenza hospitalization data

Influenza hospitalization data, the prediction target for the CDC’s FluSight challenge, are sourced from HealthData.gov’s COVID-19 Reported Patient Impact and Hospital Capacity by State Time Series. This data set includes influenza hospitalizations for every state in the US, starting 01-11-2020.

#### FluSurv-Net

The Influenza Hospitalization Surveillance Network (FluSurv-Net), part of the Respiratory Virus Hospitalization Surveillance Network (RESP-Net), conducts population-based surveillance of laboratory-confirmed influenza hospitalizations. RESP-Net includes COVID-19 (COVID-Net) and RSV (RSV-Net) surveillance networks. FluSurv-Net gathers data on laboratory-confirmed influenza-associated hospitalizations among children and adults through a network of acute care hospitals in 14 states.

We collected influenza hospitalization rates from the EIP subset of FluSurv-Net for the available states: California, Colorado, Connecticut, Georgia, Maryland, Minnesota, New Mexico, Oregon, and Tennessee. We also collected hospitalization rates for the combined network, which we use as a proxy for the United States as a whole [25].

#### ILINet

The US Outpatient Influenza-like Illness Surveillance Network (ILINet) collects data on influenza-like illness (ILI) from healthcare providers. This program is a collaboration between the CDC and various partners, including state and local health departments, hospitals, and clinics. ILINet providers submit data weekly to a central CDC repository via the internet or fax.

From ILINet, we collected unweighted ILI percentages from September 2009 to September 2020 for all states and territories of the United States, except Florida and Puerto Rico, which did not have available ILI data [26].

### Data augmentation approach

To solve the problem of limited hospitalization data, we developed a data augmentation approach which transfers information from the ILINet domain into hospitalizations. Using a standard statistical approach, we identified the best linear model to map historical ILINet data to EIP Influenza hospitalizations for California, Colorado, Connecticut, Georgia, Maryland, Minnesota, New Mexico, Oregon, and Tennessee. With this linear model, we used ILI data for all US states and regions (except Florida and Puerto Rico) to produce an extended time series of influenza hospitalizations.

More specifically, we transformed the unweighted ILI percentage to normalize it for each state using the R function bestNormalize [27]. Next, we applied a log transformation to the EIP hospitalization data to improve normality. We then produced a simple linear regression model to predict the normalized EIP values from the normalized ILI values. We tested several options, including producing a model for each of the available states. This allowed us to cluster the states without EIP data with the states that had EIP data and use the relevant state’s model to make further predictions. However, it became clear that a single model, incorporating all states with EIP data into a single linear regression model, was generally more effective than more complex approaches. After producing this single linear regression model with all available observations, we used the ILI data to produce a hospitalization time series for each state.

Finally, we fused the historical time series with the observed and recent hospitalization data–our ground-truth and prediction target data. Due to the impact of COVID-19 and the atypical appearance of the influenza time series for the 2019-20 and 2020-21 seasons, we chose to fully eliminate those seasons from our training set and analyses. Consequently, we truncated the augmented historical time series at 6/30/2019. Additionally, we retained only ground truth data starting from 07/01/2021. To maintain an uninterrupted weekly hospitalization timeseries, we added 728 days to the date index of the augmented historical time series. For most states, the final time series starts in 2012 and runs continuously to the present.

As the 2023-2024 season ended, we were able to compare our augmentation method with the ground-truth time series over the last two seasons. We generated all plots using ggplot [28].

### Forecasting approach

#### Constructing baseline forecasting models

Each horizon denotes the number of weeks ahead that we are forecasting. Thus, horizon 1 forecasts 1 week ahead, horizon 2 forecasts 2 weeks ahead, and so forth. It is important to note that we used different base models for the two seasons reported in this manuscript.

For the 2022-23 season, we primarily utilized ARIMA and regularized VAR. To construct our ARIMA model, we used the AutoARIMA method in the sktime package in Python [29, 30]. We used the fused time series and all data prior to 10/01/2022 to train the models. The model was constructed with the pipeline functionality to allow automatic power series normalization of the input data. We also constructed a separate horizon 4 model for each state, Puerto Rico, and the United States as a whole.

The second model used during the 2022-23 season is VAR. To construct this model, we used the BigVAR package. Since VAR requires the same amount of data for every covariate in the model, the limited ability to extend the time series for Florida and Puerto Rico forced us to produce two separate VAR models. One model includes all jurisdictions (including Florida and Puerto Rico) with historical data from 07/03/2021. The second VAR model excludes Florida and Puerto Rico to utilize the entire extended time series. For the model itself, we used four lags. For regularization, we selected the hierarchical lag own/other method to balance training efficiency and allow the model to prioritize its own lags over the time series of other states.

For the 2023-24 season, we constructed two base models. The first was ARIMA, produced in exactly the same manner as for the 2022-23 season. The second model was LightGBM (LGBM), implemented in the Darts Python package [31]. We constructed three separate LGBM models, each involving fully independent hyperparameter optimization. In all cases, Optuna was used to optimize model hyperparameters using a Bayesian approach. Among the LGBM models, one was optimized using 50 steps with all historical data until 06/01/2022. The second model included all data until 06/01/2022 for training along with 100 Bayesian steps. The third model included all data until 06/01/2023 along with 50 Bayesian steps for optimization. The remainder of the model specifications were identical. We used 5 as the maximum horizon number in the model because it was unclear before the season started if the CDC would require 4 or 5 horizons, and hyperparameter tuning takes approximately a week for each model. We used the following hyperparameter space for the Bayesian search.

~~~
params = {
   ‘metric’: trial.suggest_categorical(‘metric’, [‘rmse’]),
   ‘n_estimators’: trial.suggest_int(‘n_estimators’, 100, 1000, step=100),
   ‘num_leaves’: trial.suggest_int(‘num_leaves’, 8, 32, step=1),
   ‘max_depth’: trial.suggest_int(‘max_depth’, 2, 5),
   ‘learning_rate’: trial.suggest_categorical(‘learning_rate’, [0.1]),
   ‘min_child_samples’: trial.suggest_int(‘min_child_samples’, 2, 16),
   }
~~~

In addition, we use relative RMSE (rRMSE) as the loss function. rRMSE is specified as:

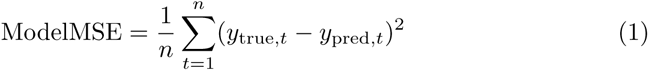

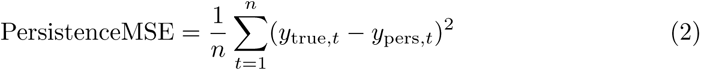

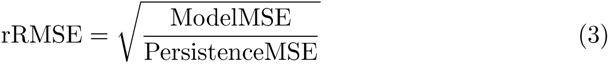

Where *y*_true*,t*_ is the actual value of the time series at time *t*, *y*_pred*,t*_ is the predicted value at time series at time *t*, and *y*_pers*,t*_ is the value of the appropriate persistence model at time *i*. Importantly, at time *t* all horizons are computed and all of the predictions are included in the loss function simultaneously. As a result, each state and the United States has a single set of hyperparameters for each of the three models that were used throughout the forecasting competition. This was a critical insight to stabilize forecasts and protect against overfitting. Notably, we use Google Cloud for hyperparameter optimization.

#### Producing final forecasts

To improve the robustness and accuracy of our forecasts, we combined each of the individual models *ŷ*^(*i*)^ into a weighted average ensemble *ỹ*. That is, at any time *t*, we assume 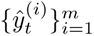 to be the set of *m* available base models. Then the ensemble prediction is as follows.

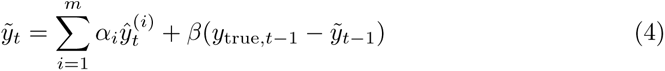

with 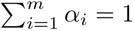 and *β ∈* [0, 1]. The weights *α_i_* were determined and adjusted weekly based on each model’s most recent historical performance, i.e., by approximately minimizing Eq. 1 over the previous 6 weeks. The ensemble also contains a bias correction term *β* based on the last observed residual of the ensemble with the target, which can adjust for systematic under- or over-prediction across the models. Because the residual can fluctuate randomly, in practice we set *β* to be the small value of 0.1 for the 2022-23 season. In the 2023-24 season, we set *β* = 0 due to the the small number of base models and the high accuracy of the LGBM models. This procedure was performed independently for each location and horizon.

#### Error analysis of FluSight models

We include error calculations for each of the top 5 models along with each of our component models for each season. Forecasts for all models included in the FluSight project are available on GitHub. The forecasts are reported probabilistically; therefore, we used the 0.5 quantile estimate for each model, prediction date, and horizon to compute RMSE and rRMSE. According to the CDC, the top 5 models for the 2022-23 season were MOBS Gleam, CMU Timeseries, PSI-DICE, MIGHTE Ensemble (our model), and CDC Ensemble. Similarly, the top 5 models for the 2023-24 season were UMass-flusion, CDC Ensemble, MIGHTE Ensemble (our model), UGA flucast-INFLAenza, and MOBS Gleam. For final display, models are arranged by the rank of their rRMSE for each horizon. It is important to note that the interpretation of horizon 1 during the 2023-24 season is challenging because the ground-truth value was available at the time of submission. Consequently, some teams reported this value directly (we chose to actually forecast it rather than include the ground-truth value). As a result, the CDC did not include that horizon in their ranking.

## Results

### Data augmentation analysis

For most locations, except Florida and Puerto Rico, we successfully extended the available historical data on influenza hospitalizations back to 2012. This extension is crucial to enable machine learning models to be trained with our proposed transfer learning approach, and helps us overcome the limitation imposed by the available sparse historical data. To produce models for extending historical hospitalizations from ILI, we first ensured that the timing and magnitude of hospitalizations and ILI were visually similar to the untransformed values (Figure 1). We subsequently tested several normalization methods and found that log-transforming EIP hospitalizations and using the bestNormalize package to perform ordered quantile normalization on the ILI data produced a very clear linear relationship in all states (Figure 2). The models for each location showed a strong connection between the two quantities after transformation (Table 1). To increase robustness, we elected to make a single combined model for all states. This final model was able to explain more than 50% of the variation in EIP hospitalizations from ILI.

**Fig 1.**
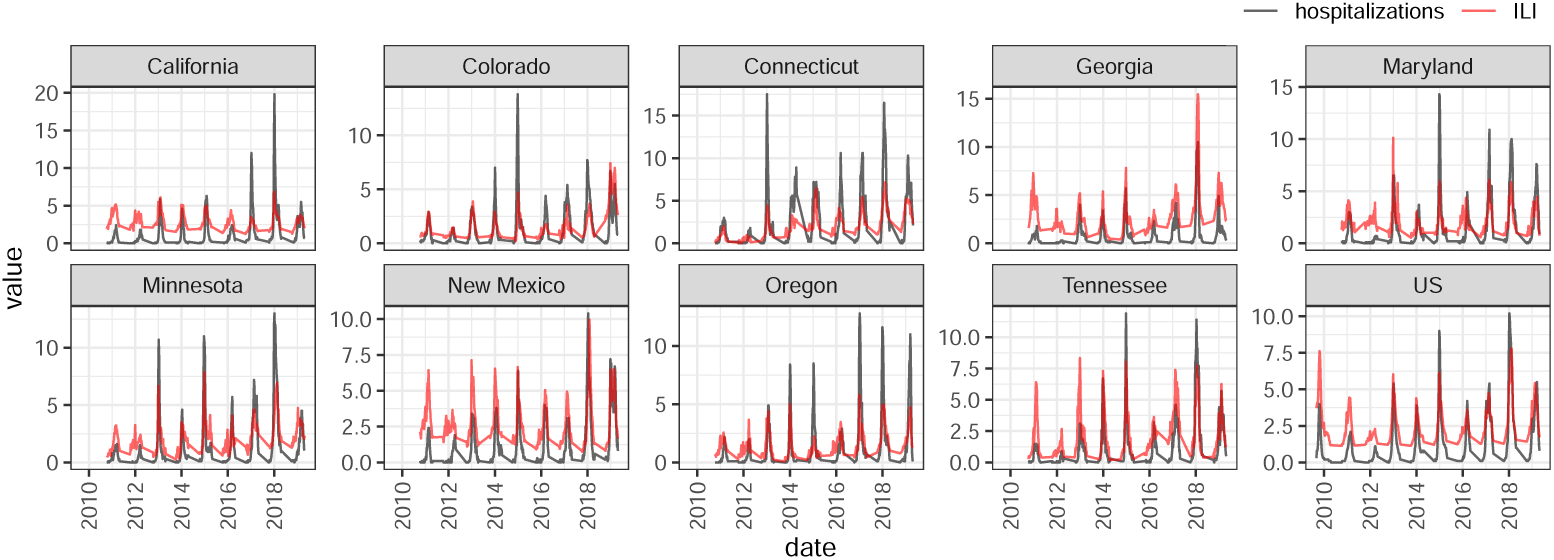
Comparison of EIP Hospitalizations and ILI across all locations with available EIP data. This figure compares the untransformed values for EIP hospitalizations and unweighted ILI percentage at all of the 8 sites with EIP values available. Based on the visualization the timing of hospitalization is increase should be well predicted from ILI while the peak may be less accurate.

**Fig 2.**
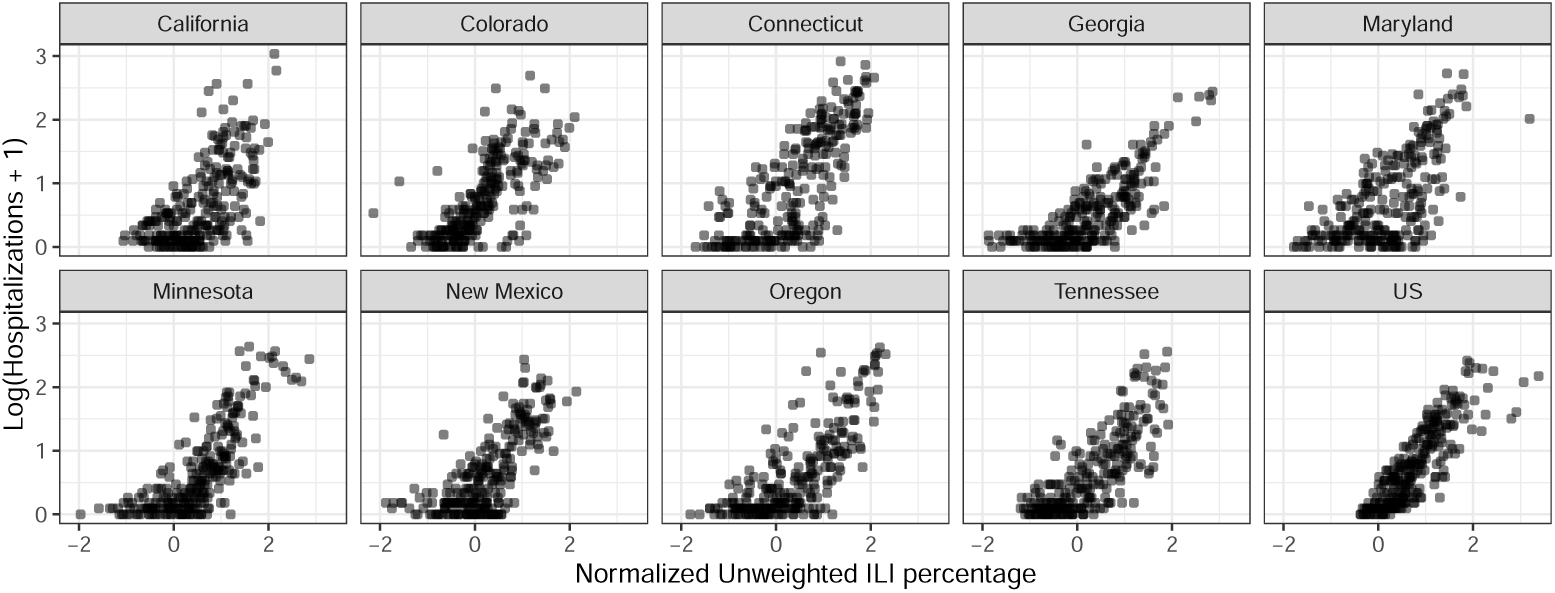
Scatter plot of log-transformed EIP Hospitalizations and normalized ILI percentage across all locations with available EIP data. This figure compares the log-transformed values for EIP hospitalizations and normalized unweighted ILI percentage at all of the 8 sites with EIP values available. Based on the visualization there should be a strong linear correlation between the two variables.

**Table 1.**
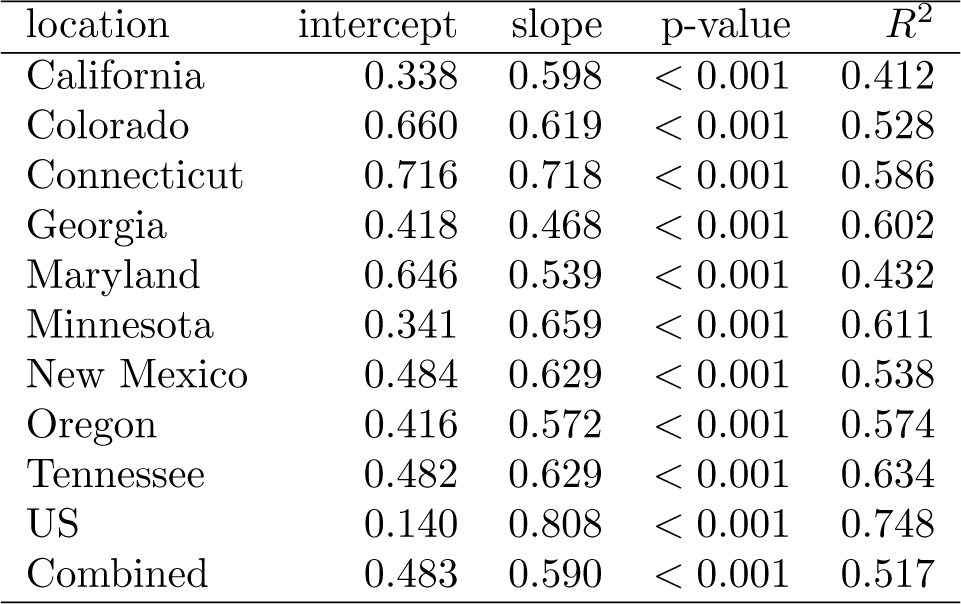
Models to Extend Historical Hospitalizations Timeseries from ILI Data. The table shows the model parameters creating by fitting log transformed EIP hospitalizations with normalized ILI unweighted percentage for all locations with EIP data. We also include the combined final model that we used to extend historical hospitalizations from ILI. Every model was highly statistically significant with the combined model explaining greater than 50% of the variation in EIP hospitalizations.

To assess the out-of-sample accuracy of our extended time series, we compared the augmented hospitalization data with actual data from the two most recent seasons (these seasons were not included in the model training). As shown in Figure 3, the augmented values are closely aligned with the actual data to capture the onset and progression of the influenza seasons. However, augmented series tend to underestimate the magnitudes of hospitalizations. This underestimation is further highlighted in the scatter plot in Figure 4, indicating a slight systematic bias in peak estimations that may lead to suboptimal peak predictions when producing forecasts on the hospitalization time series.

**Fig 3.**
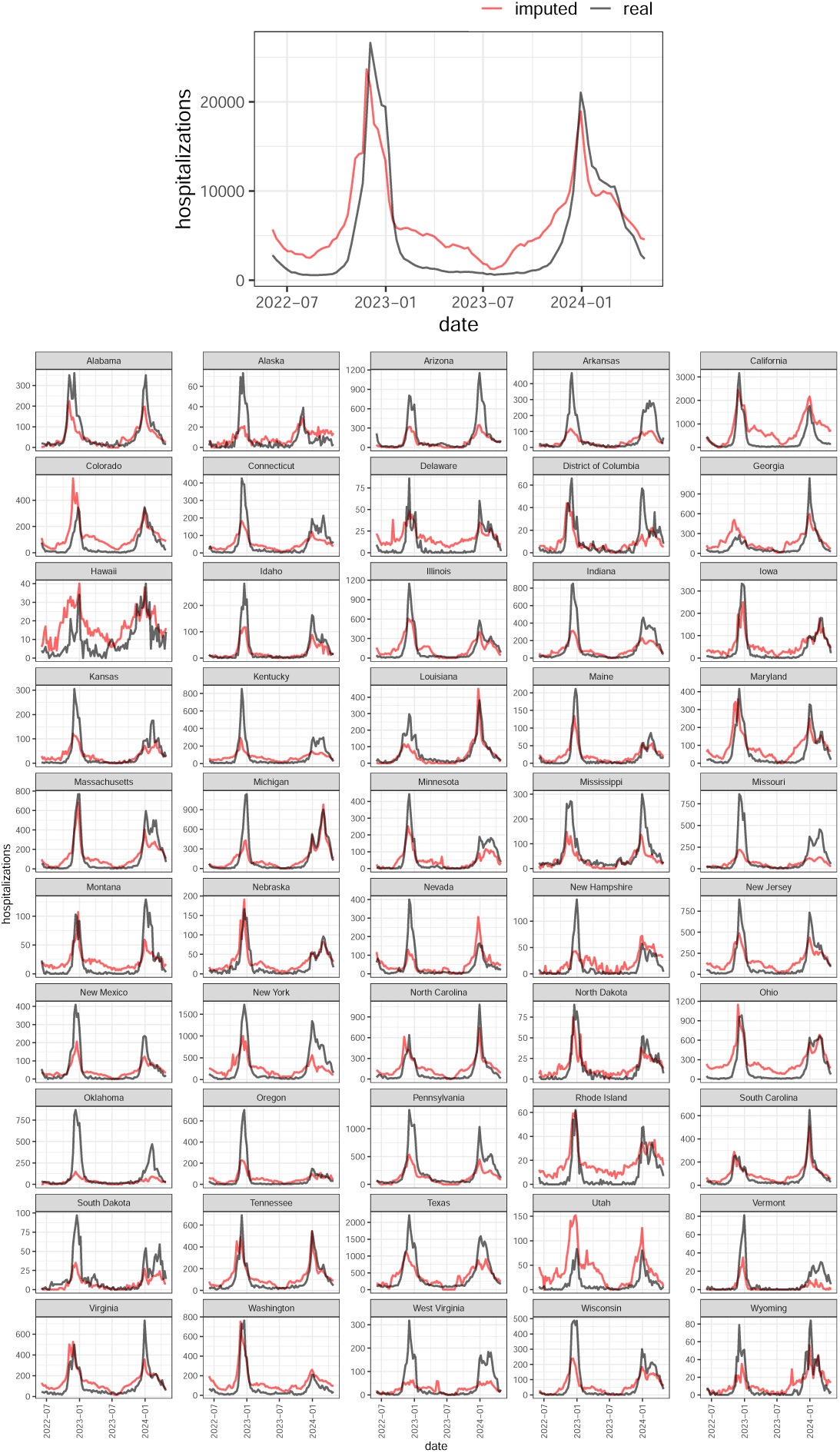
Comparison of Augmented Hospitalizations and Actual Hospitalizations Across All States and Nationwide. The figure shows the augmented hospitalizations (not forecasts) against the actual (ground truth) hospitalization time series for all states and the United States as a whole. The top figure represents the United States as a whole. The comparison evaluates the model’s ability to accurately recapitulate the timing of influenza season onset and peak hospitalization periods. While the augmented values closely align with the actual data in terms of timing, indicating the model’s effectiveness in capturing seasonal trends, there is a notable discrepancy in peak magnitude estimation.

**Fig 4.**
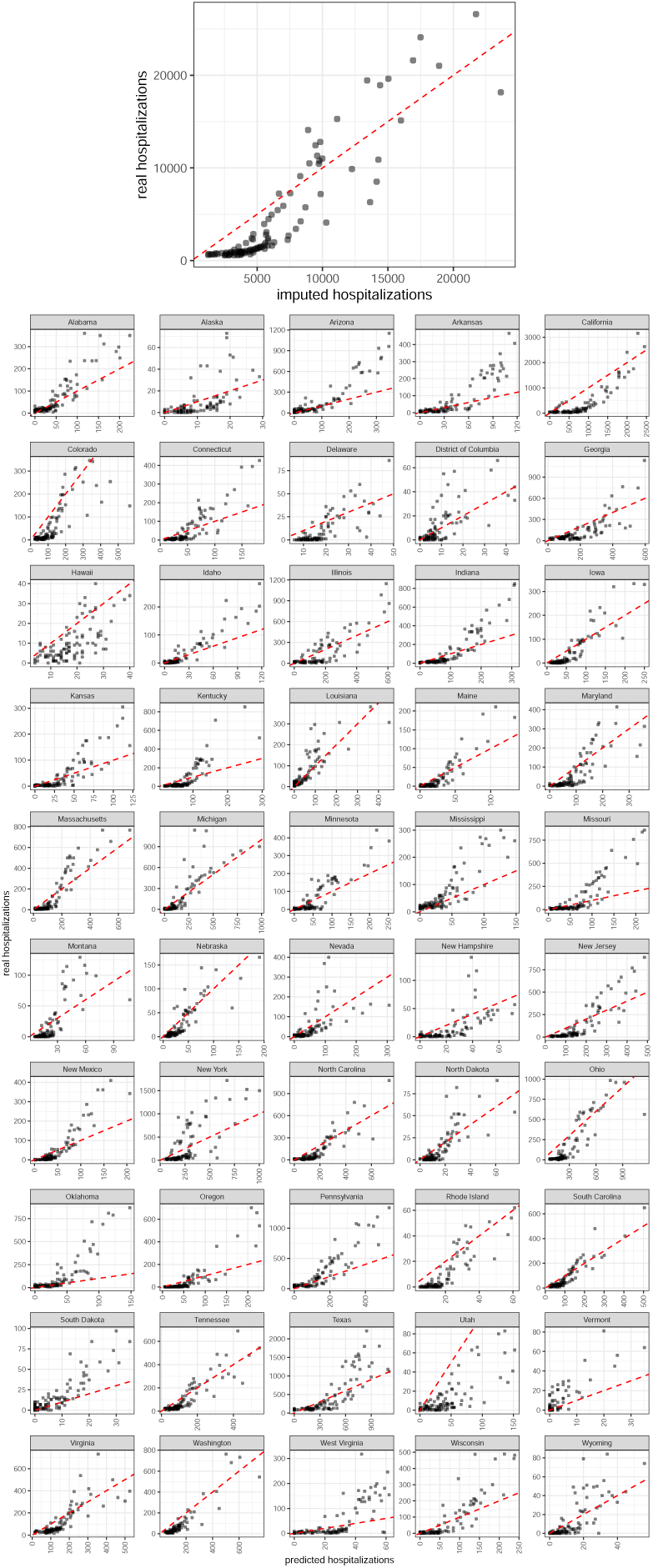
Scatter Plot Analysis of Augmented Hospitalizations Versus Actual Hospitalizations for the 2022-23 and 2023-24 Seasons. This figure provides a scatter plot comparison between the augmented hospitalizations (not forecasts) and actual (ground truth) hospitalizations for the influenza seasons of 2022-23 and 2023-24 for all states and the United States as a whole; the top figure is the United States as a whole.The dashed red line, representing a perfect match (slope = 1, intercept = 0), serves as a reference to evaluate the accuracy of our predictions. The general alignment of augmented values with the actual data suggests that the forecasting models serve as an effective proxy for real-time series analysis across the examined periods. However, augmented values near peak hospitalization periods exhibit a consistent trend of slight overestimation, highlighting a potential area for refinement in model accuracy. The visual representation underscores the overall reliability of the predictive models to augmented hospitalizations while also identifying specific intervals where further model adjustments could enhance forecasting precision.

Despite these discrepancies, a quantitative analysis using a combined linear fit between augmented and actual hospitalizations results in an adjusted *R*^2^ value of 0.8, with an intercept of zero and a slope of 1.1. This indicates a strong correlation and good accuracy for the extended time series, particularly in terms of temporal trends and overall magnitude, although peak values remain an exception.

Given the close timing alignment and reasonable accuracy in overall magnitude (excluding peak values), we utilized these augmented data sets to develop our forecasting models.

### Forecasting analysis

We generate strictly out-of-sample influenza hospitalization forecasts for each state in the United States using our proposed ensemble methodologies. The evaluation periods aligned with the official influenza seasons (as defined by the CDC) between October 17, 2022, through May 17, 2023 (22-23 season) and October 11, 2023, through May 1, 2024 (23-24 season). Specifically, for every week and in every state, we generated a forecast up to 4 weeks in advance for 2022-23 and up to 5 weeks in advance for 2023-24.

#### Comparing models with and without augmented data

To validate the impact of our extension method, we retrospectively compared the performance of ARIMA models trained with and without augmented data for both the 2022-2023 and 2023-2024 seasons. These results are shown in Tables 2 and 3.

**Table 2.**
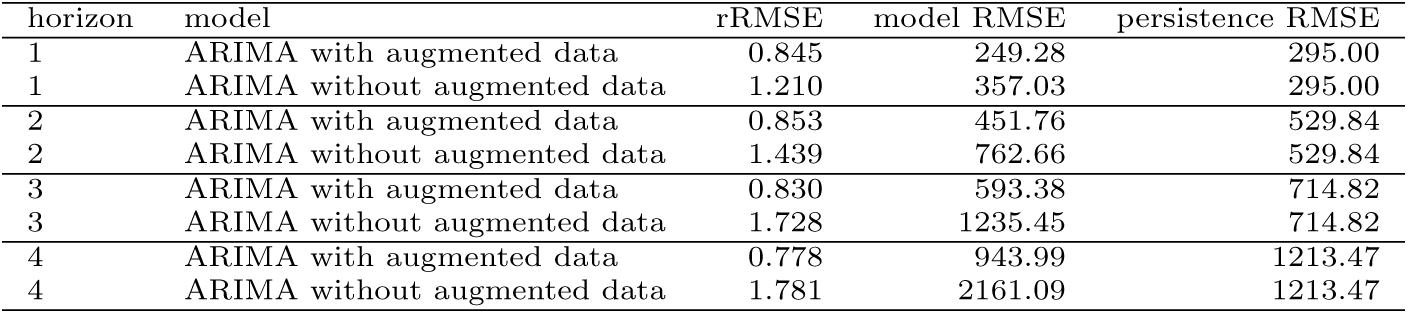
Comparative Performance of ARIMA with and without augmented data for the 2022-23 season. This table shows the forecast error using an ARIMA model with and without augmented data for each horizon during the 2022-23 season. Each horizon is ordered by rRMSE which is defined as the ratio of model RMSE to persistence RMSE. ARIMA with augmented data significantly outperforms ARIMA without augmented data at all horizons.

| horizon | model | rRMSE | model RMSE | persistence RMSE |
| --- | --- | --- | --- | --- |
| 1 | ARIMA with augmented data | 0.845 | 249.28 | 295.00 |
| 1 | ARIMA without augmented data | 1.210 | 357.03 | 295.00 |
| 2 | ARIMA with augmented data | 0.853 | 451.76 | 529.84 |
| 2 | ARIMA without augmented data | 1.439 | 762.66 | 529.84 |
| 3 | ARIMA with augmented data | 0.830 | 593.38 | 714.82 |
| 3 | ARIMA without augmented data | 1.728 | 1235.45 | 714.82 |
| 4 | ARIMA with augmented data | 0.778 | 943.99 | 1213.47 |
| 4 | ARIMA without augmented data | 1.781 | 2161.09 | 1213.47 |

**Table 3.** Comparative Performance of ARIMA with and without augmented data for the 2023-24 season. This table shows the forecast error using an ARIMA model with and without augmented data for each horizon during the 2023-24 season. Each horizon is ordered by rRMSE which is defined as the ratio of model RMSE to persistence RMSE. ARIMA with augmented data significantly outperforms ARIMA without augmented data at all horizons.

| horizon | model | rRMSE | model RMSE | persistence RMSE |
| --- | --- | --- | --- | --- |
| 1 | ARIMA with augmented data | 0.857 | 233.72 | 272.58 |
| 1 | ARIMA without augmented data | 1.294 | 352.59 | 272.58 |
| 2 | ARIMA with augmented data | 0.887 | 439.89 | 495.70 |
| 2 | ARIMA without augmented data | 1.578 | 782.01 | 495.70 |
| 3 | ARIMA with augmented data | 0.882 | 589.34 | 668.37 |
| 3 | ARIMA without augmented data | 2.020 | 1350.42 | 668.37 |
| 4 | ARIMA with augmented data | 0.861 | 693.60 | 805.36 |
| 4 | ARIMA without augmented data | 2.396 | 1929.39 | 805.36 |

For both seasons, ARIMA models with augmented data consistently reduced the baseline error (compared to persistence) across all forecast horizons. Specifically, our models achieved error reductions ranging from 11% (e.g., 2023-2024 season, horizon 3) to 22% (e.g. 2022-2023 season, horizon 4). In contrast, models trained on the original dataset without augmentation did not manage to achieve an RMSE below the persistence baseline. The lowest error achieved by ARIMA without augmented data was approximately 21% above the baseline error.

#### Utilizing augmented data to make prospective forecasts

It is important to note that, given the prospective nature of this forecasting challenge, not all models consistently submitted forecasts for each location and each week during both seasons, making performance comparison more difficult. To address this, we include a relative RMSE metric (rRMSE), which involves normalizing a model’s RMSE score by the RMSE score of the persistence baseline (a naive model where the hospitalization forecasts for each horizon are the most recent observed activity at time *t*, or *y_t_* + *k* = *y_t_*). In other words, we compare the performance of our models by evaluating their ability to reduce the RMSE compared to the persistence model on the dates the models were submitted during the challenge.

The forecasting analysis was conducted in two main phases: constructing baseline models and integrating them into an ensemble. We evaluated various types of model, including exponential smoothing, ARIMA, VAR, regularized VAR, custom linear models with external covariates like ARGO, and gradient-boosted machines. For the 2022-23 season, we were asked to provide forecasts within a week of notification (ultimately, we were only able to provide a forecast starting in the second week that season), limiting the time for hyperparameter tuning. In contrast, the 2023-24 season allowed significant lead time for training, although the operational timeline from data release to submission was within 12 hours, necessitating a strategic selection of base models.

For the 2022-23 season, we used simpler models such as ARIMA and VAR that required minimal tuning. For the 2023-24 season, we prioritized LGBM models due to their strong out-of-sample performance in the prior season across all forecasting horizons. Each LGBM model was initialized with unique random seeds for ensemble integration. Due to the lack of long-term ILI data for Florida and Puerto Rico, preventing the creation of augmented datasets for these areas, ARIMA models were incorporated to ensure a forecast for every site.

During the 2022-23 season, the ARIMA base model had the lowest error in terms of our models (consistently reducing the error compared to the persistence baseline by up to 23% at horizon 4, and similarly across the rest of the horizons), particularly for the United States as a whole (Table 4). VAR with augmented data outperformed the naive persistence model in more locations than ARIMA. Both VAR with augmented data and ARIMA exceeded the performance of the persistence model across all horizons. VAR without augmented data performed similarly to persistence (rRMSE was 1.056, 0.993, 0.943, and 1.049 for horizons 1 through 4), but showed degraded performance with increasing horizons. Furthermore, VAR with augmented data outperformed VAR without augmented data at all horizons (rRMSE of 0.982, 0.936, 0.921, and 0.872 for horizons 1 through 4 with augmented data versus 1.056, 0.993, 0.943, and 1.049 for horizons 1 through 4 without augmented data). Comparing our MIGHTE Ensemble to the top FluSight models, we performed well for near-term forecasts, but showed declining performance at more distant horizons. Our ensemble methodology consistently reduced the error across all horizons in comparison to the persistence model, with scores of 0.837, 0.868, 0.911 and 0.876 rRMSE, respectively.

**Table 4.** Comparative Performance of Forecasting Models Across Horizons for the 2022-23 Season. This table presents the forecasting errors for each of our base models along with the top 5 models in the FluSight collaborative and a naive persistence model for reference. rRMSE is the ratio of the model RMSE to the persistence RMSE. Models are arranged from best rRMSE to worst rRMSE within each horizon category. Notably, the model that does not include augmented data performs significantly worse than the models with augmented data, and often performs worse than the persistence model. Note: as all forecasts were prospective, persistence models vary slightly due to missing a small number of weekly forecasts as a result of time constraints and/or differing locations included.

| horizon | model | rRMSE | model RMSE | persistence RMSE |
| --- | --- | --- | --- | --- |
| 1 | PSI-DICE | 0.753 | 310.00 | 411.00 |
| 1 | CDC Ensemble | 0.808 | 332.00 | 411.00 |
| 1 | **MIGHTE Ensemble** | 0.837 | 350.00 | 418.00 |
| 1 | ARIMA | 0.863 | 359.68 | 416.59 |
| 1 | VAR augmented data | 0.982 | 408.01 | 415.54 |
| 1 | CMU Timeseries | 0.999 | 411.00 | 411.00 |
| 1 | MOBS Gleam | 1.040 | 427.00 | 418.00 |
| 1 | VAR no augmented data | 1.056 | 414.34 | 392.30 |
| 2 | PSI-DICE | 0.816 | 598.00 | 733.00 |
| 2 | ARIMA | 0.839 | 618.34 | 737.36 |
| 2 | MOBS Gleam | 0.853 | 625.00 | 746.00 |
| 2 | **MIGHTE Ensemble** | 0.868 | 648.00 | 746.00 |
| 2 | CDC Ensemble | 0.889 | 652.00 | 733.00 |
| 2 | VAR augmented data | 0.936 | 697.16 | 744.81 |
| 2 | VAR no augmented data | 0.993 | 583.88 | 588.16 |
| 2 | CMU Timeseries | 1.010 | 742.00 | 746.00 |
| 3 | MOBS Gleam | 0.665 | 658.00 | 989.00 |
| 3 | PSI-DICE | 0.753 | 745.00 | 989.00 |
| 3 | CMU Timeseries | 0.757 | 749.00 | 989.00 |
| 3 | ARIMA | 0.805 | 797.09 | 990.55 |
| 3 | CDC Ensemble | 0.857 | 847.00 | 989.00 |
| 3 | **MIGHTE Ensemble** | 0.911 | 916.00 | 1006.00 |
| 3 | VAR augmented data | 0.921 | 932.17 | 1012.27 |
| 3 | VAR no augmented data | 0.943 | 743.40 | 788.55 |
| 4 | MOBS Gleam | 0.490 | 595.00 | 1214.00 |
| 4 | PSI-DICE | 0.631 | 767.00 | 1214.00 |
| 4 | CMU Timeseries | 0.692 | 840.00 | 1214.00 |
| 4 | ARIMA | 0.777 | 944.45 | 1214.96 |
| 4 | CDC Ensemble | 0.839 | 1019.00 | 1214.00 |
| 4 | VAR augmented data | 0.872 | 1092.48 | 1252.97 |
| 4 | **MIGHTE Ensemble** | 0.876 | 1081.00 | 1235.00 |
| 4 | VAR no augmented data | 1.049 | 1045.43 | 996.13 |

In the 2023-24 season, at least one LGBM model showed superior predictive accuracy compared to the naive persistence model across all horizons. At horizon 1, as in 2022-23, the ARIMA model showed respectable accuracy. The performance metrics of the individual models, outlined in Table 5, varied across horizons. A LGBM model using data up to June 2023 provided the most accurate forecasts for horizons 1 through 3 (with values of 0.969 and 0.706 rRMSE, respectively), while a LGBM model using data up to June 1, 2022, was most effective for the fourth horizon (0.699 rRMSE). Comparing our MIGHTE Ensemble to other top FluSight models, our ensemble showed improved relative accuracy as the horizon number increased (down to 0.719 rRMSE in horizon 4). The small magnitude of error growth across horizons likely resulted from implementing a loss function that included forecasts for all horizons simultaneously, enhancing model robustness by balancing near- and intermediate-term costs. Notably, the model that consistently outperformed our ensemble also used historical augmentation and gradient-boosted models.

**Table 5.**
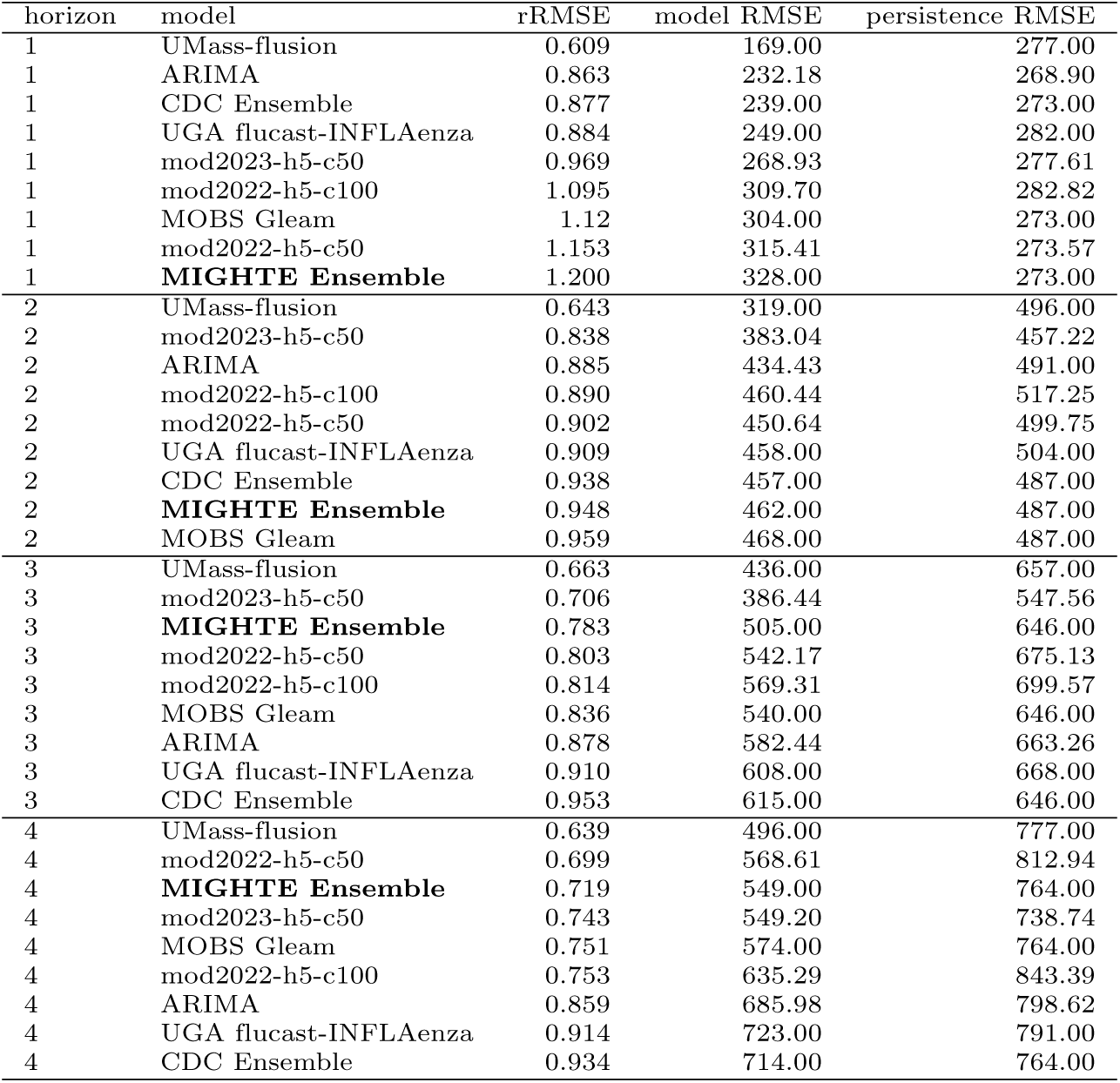
Comparative Performance of Forecasting Models Across Horizons for the 2023-24 Season. This table presents the forecasting errors for each of our base models along with the top 5 models in the FluSight collaborative and a naive persistence model for reference. rRMSE is the ratio of the model RMSE to the persistence RMSE. Models are arranged from best rRMSE to worst rRMSE within each horizon category. Notably, LGBM models demonstrate substantial accuracy beyond the initial horizon, with minimal degradation in performance observed up to horizon 4. Note: as all forecasts were prospective, persistence models vary slightly due to missing a small number of weekly forecasts as a result of time constraints and/or differing locations included.

| horizon | model | rRMSE | model RMSE | persistence RMSE |
| --- | --- | --- | --- | --- |
| 1 | UMass-flusion | 0.609 | 169.00 | 277.00 |
| 1 | ARIMA | 0.863 | 232.18 | 268.90 |
| 1 | CDC Ensemble | 0.877 | 239.00 | 273.00 |
| 1 | UGA flucast-INFLAenza | 0.884 | 249.00 | 282.00 |
| 1 | mod2023-h5-c50 | 0.969 | 268.93 | 277.61 |
| 1 | mod2022-h5-c100 | 1.095 | 309.70 | 282.82 |
| 1 | MOBS Gleam | 1.12 | 304.00 | 273.00 |
| 1 | mod2022-h5-c50 | 1.153 | 315.41 | 273.57 |
| 1 | **MIGHTE Ensemble** | 1.200 | 328.00 | 273.00 |
| 2 | UMass-flusion | 0.643 | 319.00 | 496.00 |
| 2 | mod2023-h5-c50 | 0.838 | 383.04 | 457.22 |
| 2 | ARIMA | 0.885 | 434.43 | 491.00 |
| 2 | mod2022-h5-c100 | 0.890 | 460.44 | 517.25 |
| 2 | mod2022-h5-c50 | 0.902 | 450.64 | 499.75 |
| 2 | UGA flucast-INFLAenza | 0.909 | 458.00 | 504.00 |
| 2 | CDC Ensemble | 0.938 | 457.00 | 487.00 |
| 2 | **MIGHTE Ensemble** | 0.948 | 462.00 | 487.00 |
| 2 | MOBS Gleam | 0.959 | 468.00 | 487.00 |
| 3 | UMass-flusion | 0.663 | 436.00 | 657.00 |
| 3 | mod2023-h5-c50 | 0.706 | 386.44 | 547.56 |
| 3 | **MIGHTE Ensemble** | 0.783 | 505.00 | 646.00 |
| 3 | mod2022-h5-c50 | 0.803 | 542.17 | 675.13 |
| 3 | mod2022-h5-c100 | 0.814 | 569.31 | 699.57 |
| 3 | MOBS Gleam | 0.836 | 540.00 | 646.00 |
| 3 | ARIMA | 0.878 | 582.44 | 663.26 |
| 3 | UGA flucast-INFLAenza | 0.910 | 608.00 | 668.00 |
| 3 | CDC Ensemble | 0.953 | 615.00 | 646.00 |
| 4 | UMass-flusion | 0.639 | 496.00 | 777.00 |
| 4 | mod2022-h5-c50 | 0.699 | 568.61 | 812.94 |
| 4 | **MIGHTE Ensemble** | 0.719 | 549.00 | 764.00 |
| 4 | mod2023-h5-c50 | 0.743 | 549.20 | 738.74 |
| 4 | MOBS Gleam | 0.751 | 574.00 | 764.00 |
| 4 | mod2022-h5-c100 | 0.753 | 635.29 | 843.39 |
| 4 | ARIMA | 0.859 | 685.98 | 798.62 |
| 4 | UGA flucast-INFLAenza | 0.914 | 723.00 | 791.00 |
| 4 | CDC Ensemble | 0.934 | 714.00 | 764.00 |

For detailed tracings of all models compared to ground truth, see supplementary figures.

## Discussion

In this study, we developed and evaluated an innovative approach to forecasting influenza hospitalizations by combining data augmentation, statistical models, machine learning models, and ensembling techniques. This approach overcomes the challenges of implementing advanced machine learning models to forecast influenza hospitalizations and provides additional data for training traditional statistical models. Using a simple transfer learning method with similar epidemic curves, we first trained models to predict the shape and timing of influenza-like illnesses (ILI), and then used these (sometimes fine-tuned) models to produce flu hospitalization forecasts.

Our forecasts were rigorously validated over two seasons in the CDC FluSight challenge, achieving fourth place in 2022-23 and second place in 2023-24 among 20 teams, outperforming the aggregated FluSight ensemble in one season and tying it in the other. Our findings emphasize the crucial role of data quality and availability in modeling infectious diseases.

The data augmentation strategy we used to extend the historical data set is vital to overcome sparse data limitations and training machine learning models. Specifically, we were only able to train a LightGBM due to the availability of augmented data. The baseline models that were trained with the augmented data beat virtually all of the other models in the FluSight consortium. Additional to our analysis about the impact of imputation, which showed a clear distinction between the error of ARIMA being trained using our extended dataset and the original dataset, there is clear evidence in the 2022-23 season that the augmented data alone were able to improve forecasts. In that season, we implemented regularized VAR models with and without augmented data, and VAR with augmented data consistently outperformed VAR without augmented data. Furthermore, according to the model metadata available on the public repository, the single model that consistently beat our model during the 2023-24 season also used augmented data along with a gradient-boosted machine. This finding would suggest that augmented data coupled with advanced machine learning models may offer a path forward in other cases where data sparsity presents a significant challenge.

It is important to note that our method for augmenting historical hospitalizations is far from perfect, especially in estimating peak hospitalization volumes, where it seems systematically biased to underpredict hospitalizations. This highlights the need for ongoing refinement of cross-domain data augmentation techniques and the integration of additional data sources to enhance model accuracy. Future research should focus on several key areas. First, exploring advanced augmentation methods and incorporating diverse data types could better approximate true hospitalization curves. Second, integrating real-time data streams, such as search query trends, social media analytics, or viral testing data, can improve the timeliness and accuracy of forecasts. Third, while our models are successful in influenza forecasting, their generalizability to other infectious diseases requires further investigation. Fourth, implementing additional models with loss functions that include multiple horizons, locations, and weighted components could improve robustness in cases where data sparsity is an issue. Additionally, recent advances in transformer models with transfer learning approaches similar to ours suggest that fine-tuning existing time series foundation models for respiratory virus specialization could significantly advance the field. Such research could greatly expand the applicability of machine learning techniques in public health forecasting.

In summary, our research has significant practical implications. Accurate forecasting of influenza hospitalizations enhances public health preparedness, informs resource allocation, and mitigates the impact of influenza epidemics on communities. Our work adds to the growing body of literature on machine learning in epidemiology, showcasing the potential of transfer learning to improve models for complex forecasting challenges. While promising, our approach has limitations and serves as a foundation for further research to refine and expand predictive models in public health. The ongoing integration of epidemiology and machine learning offers great potential for advancing our ability to predict and respond to infectious disease outbreaks.

## Data Availability

The data are publicly available at:
https://gis.cdc.gov/grasp/fluview/main.html
https://gis.cdc.gov/GRASP/Fluview/FluHospRates.html
https://github.com/cdcepi/FluSight-forecast-hub

## Supplementary Figures

**Fig S1.**
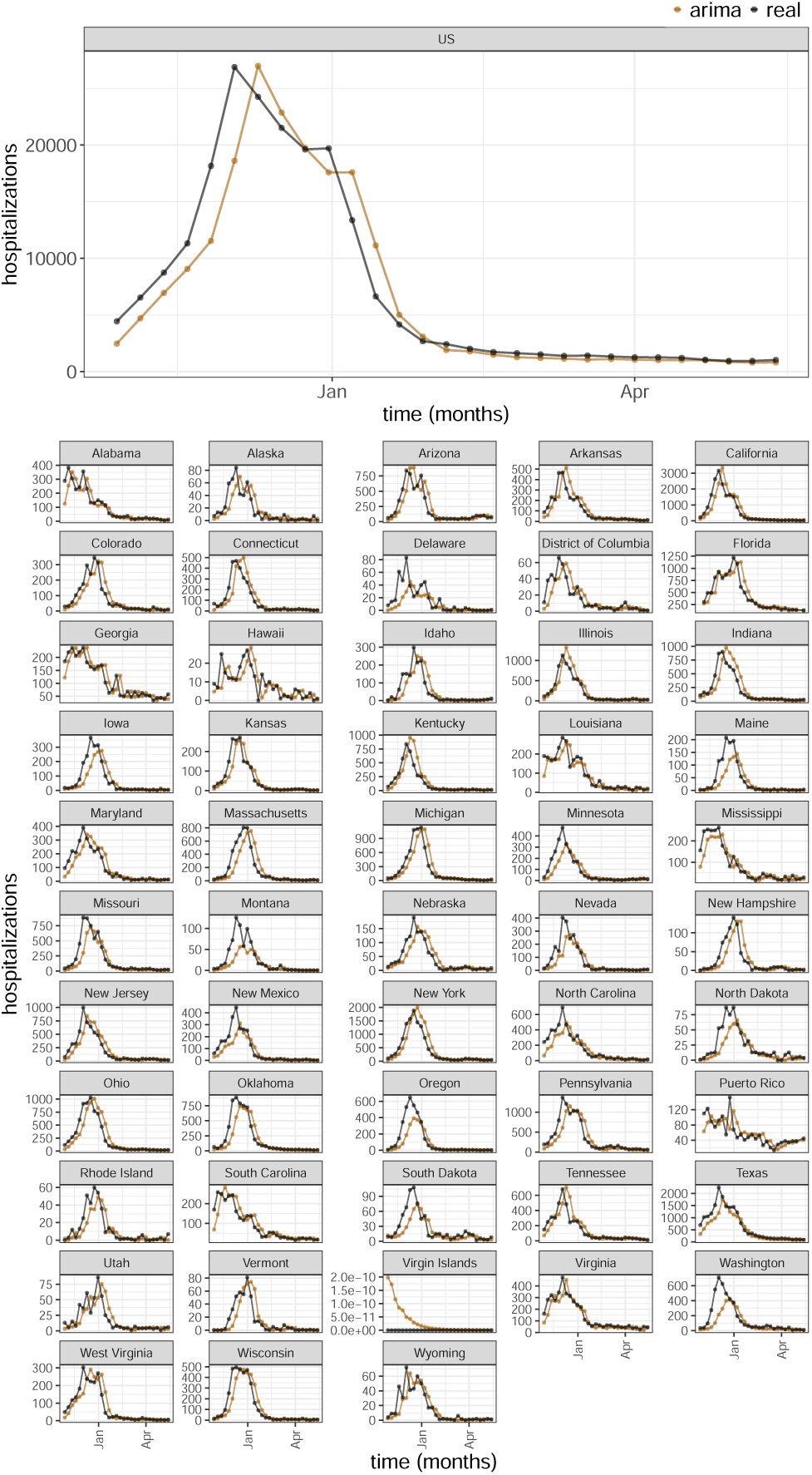
Comparison of ARIMA Model Forecasts with Ground Truth Data at Horizon 1 for the 2022-23 season. This figure illustrates the forecasting performance of the ARIMA model for the first prediction horizon (1 week ahead) relative to the actual observed hospitalizations (ground truth).

**Fig S2.**
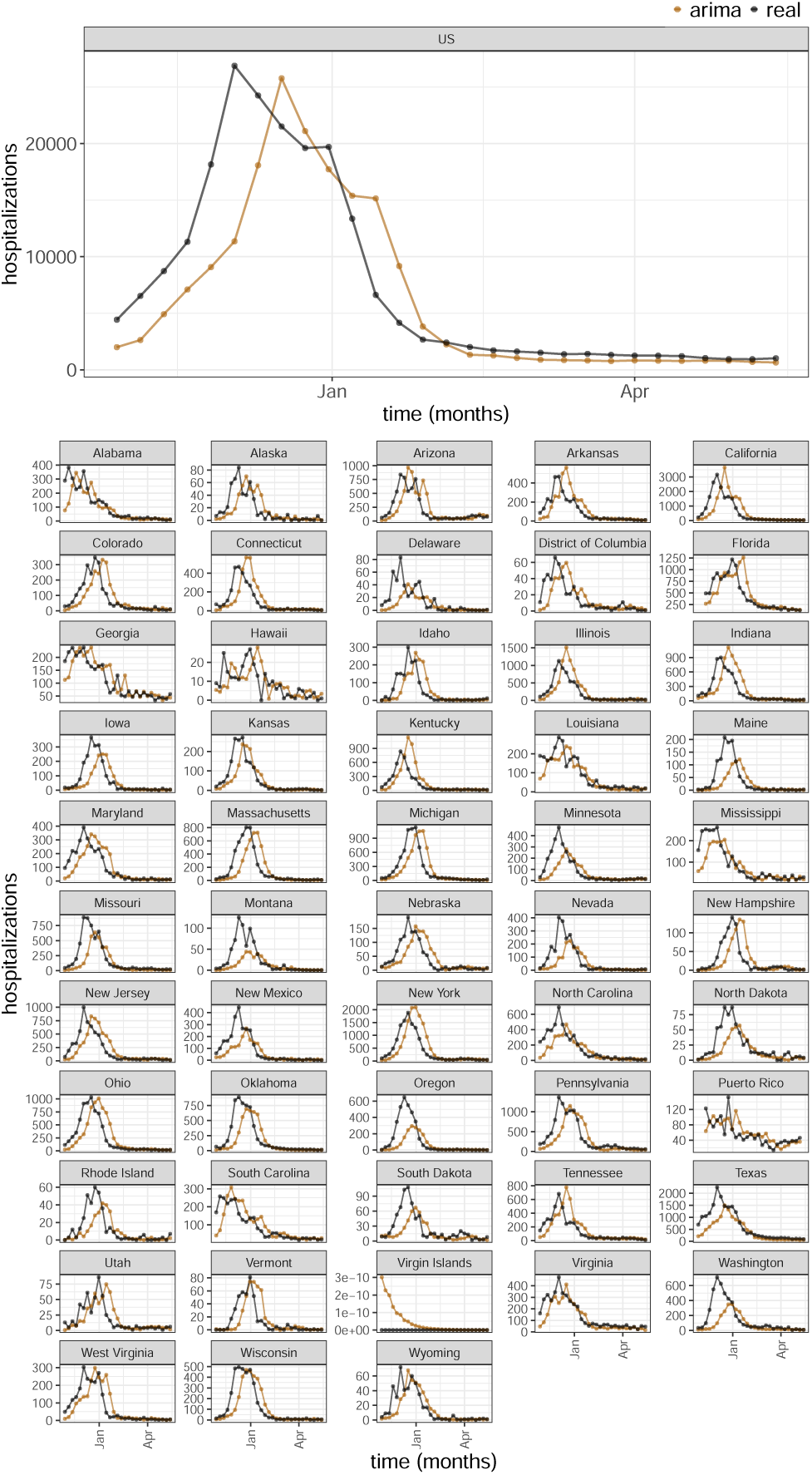
Comparison of ARIMA Model Forecasts with Ground Truth Data at Horizon 2 for the 2022-23 season. This figure illustrates the forecasting performance of the ARIMA model for the first prediction horizon (2 weeks ahead) relative to the actual observed hospitalizations (ground truth).

**Fig S3.**
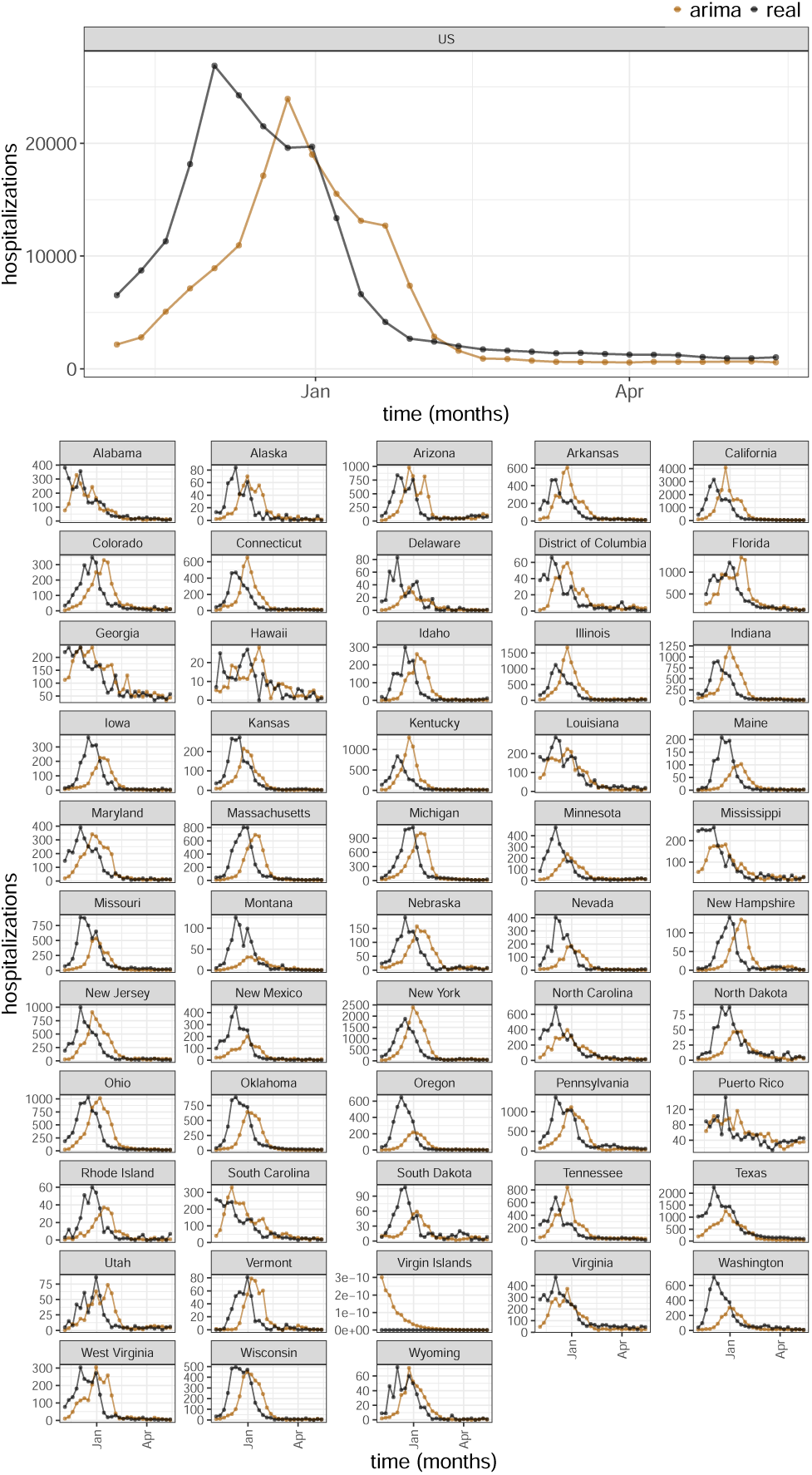
Comparison of ARIMA Model Forecasts with Ground Truth Data at Horizon 3 for the 2022-23 season. This figure illustrates the forecasting performance of the ARIMA model for the first prediction horizon (3 weeks ahead) relative to the actual observed hospitalizations (ground truth).

**Fig S4.**
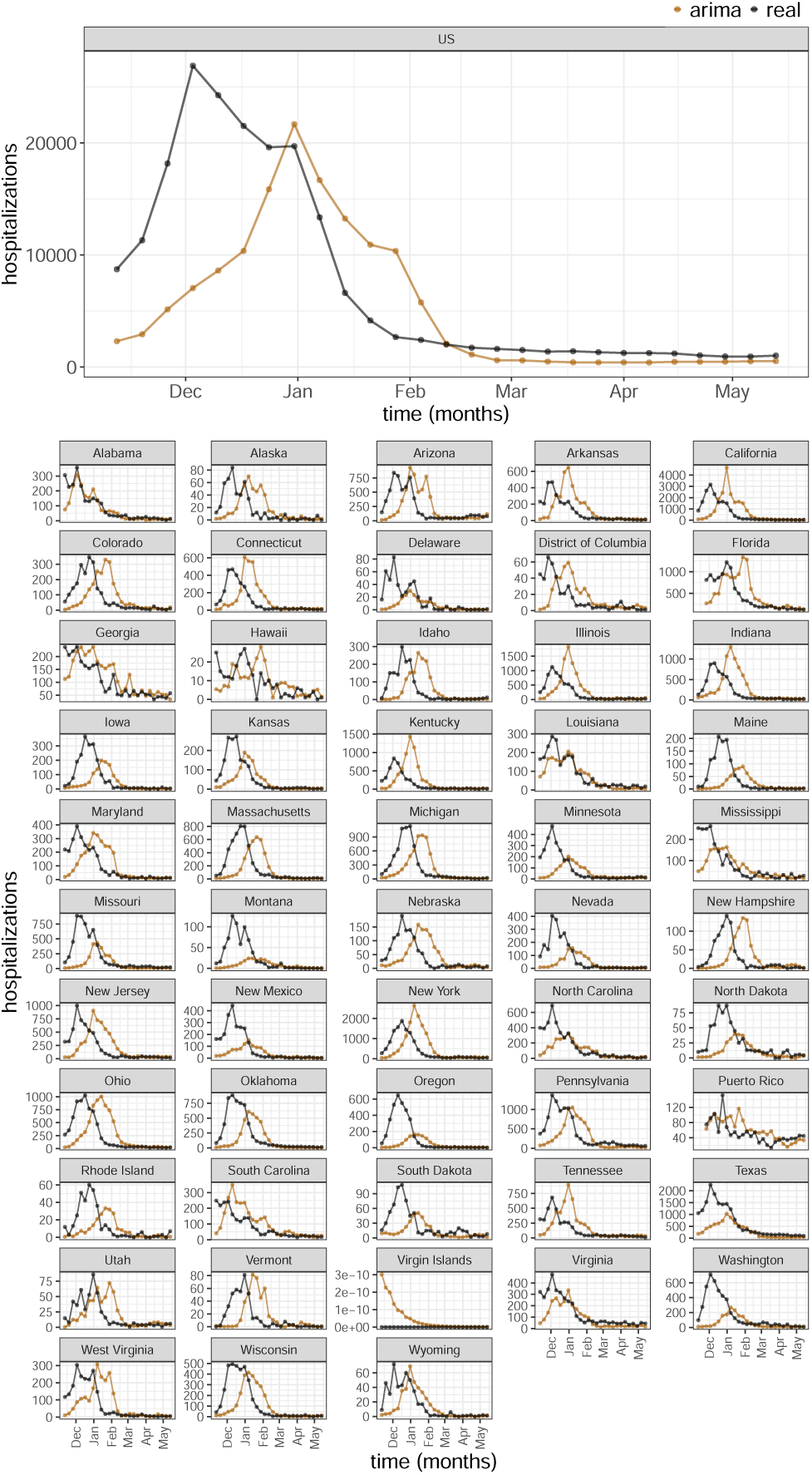
Comparison of ARIMA Model Forecasts with Ground Truth Data at Horizon 4 for the 2022-23 season. This figure illustrates the forecasting performance of the ARIMA model for the first prediction horizon (4 weeks ahead) relative to the actual observed hospitalizations (ground truth).

**Fig S5.**
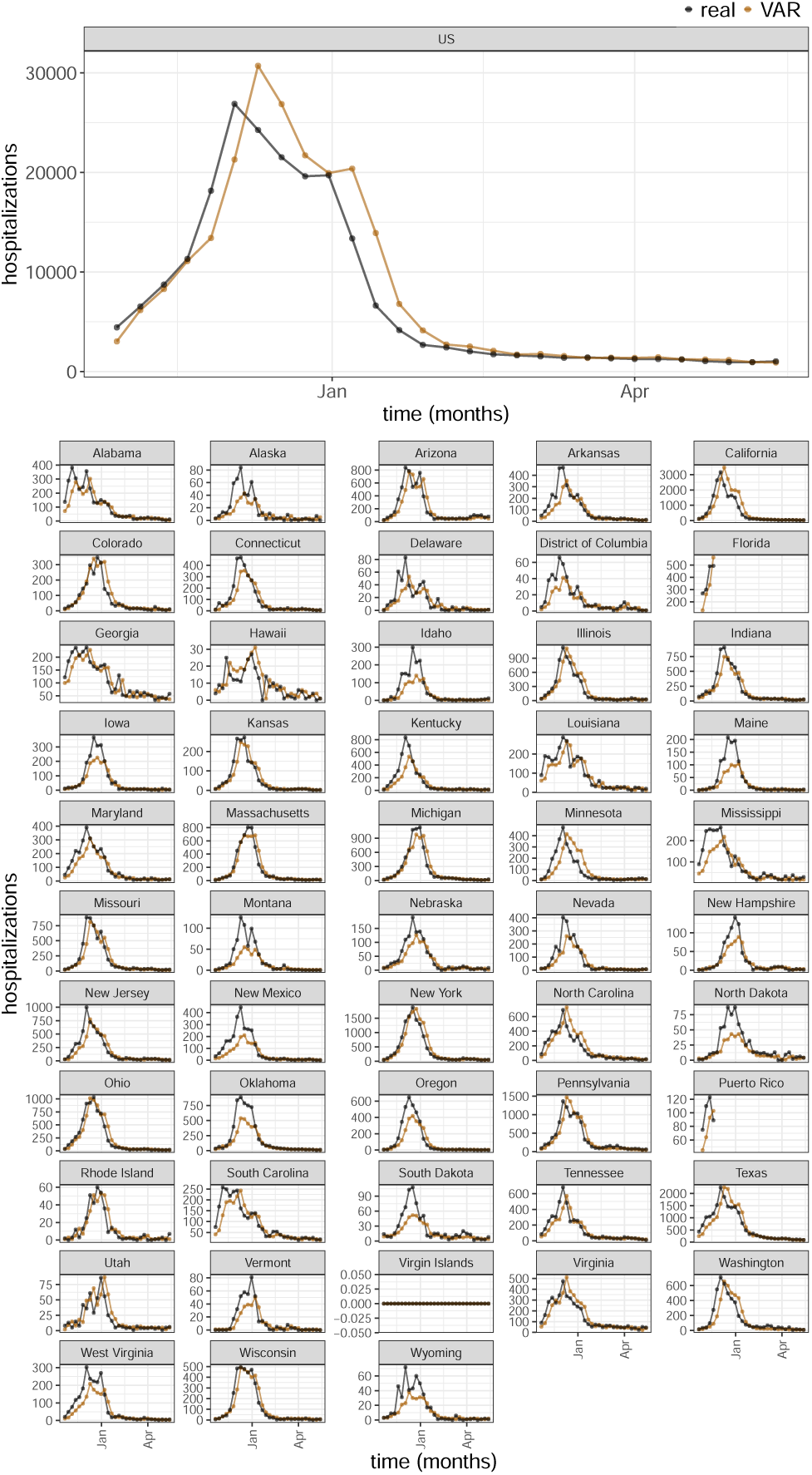
Comparison of ARIMA Model Forecasts with Ground Truth Data at Horizon 1 for the 2022-23 season. This figure illustrates the forecasting performance of the regularized VAR model for the first prediction horizon (1 week ahead) relative to the actual observed hospitalizations (ground truth).

**Fig S6.**
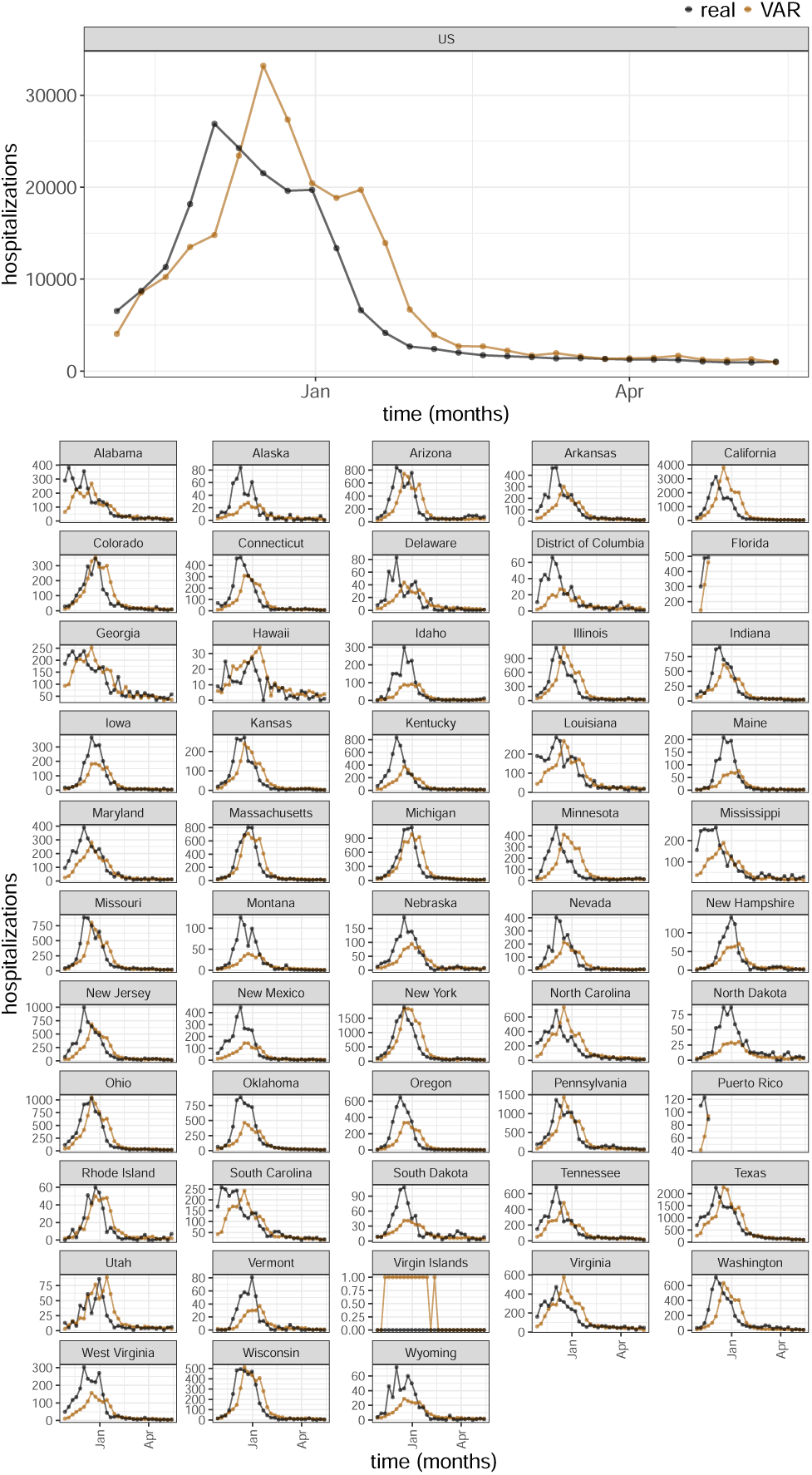
Comparison of VAR Model Forecasts with Ground Truth Data at Horizon 2 for the 2022-23 season. This figure illustrates the forecasting performance of the regularized VAR model for the first prediction horizon (2 weeks ahead) relative to the actual observed hospitalizations (ground truth).

**Fig S7.**
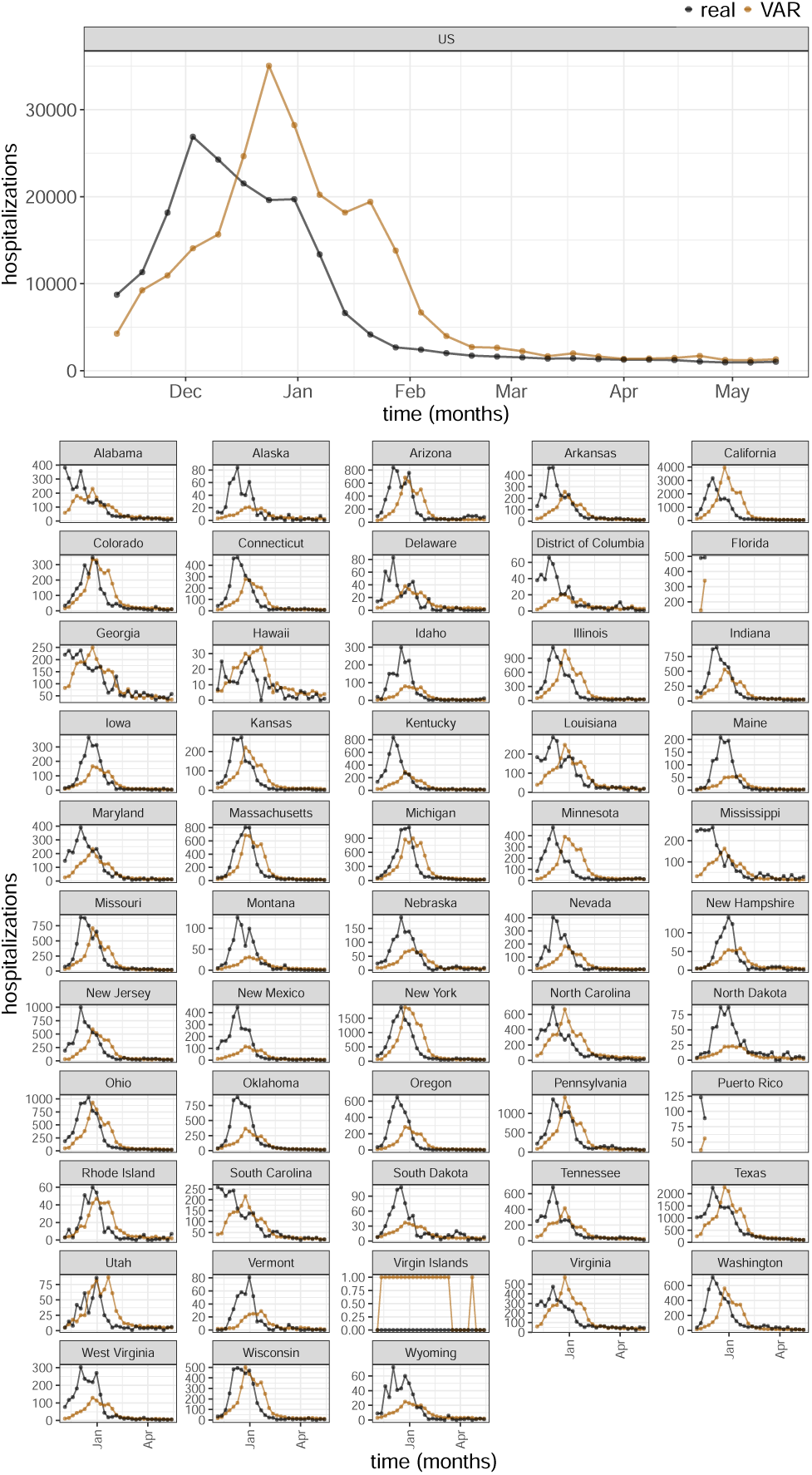
Comparison of VAR Model Forecasts with Ground Truth Data at Horizon 3 for the 2022-23 season. This figure illustrates the forecasting performance of of the regularized VAR model for the first prediction horizon (3 weeks ahead) relative to the actual observed hospitalizations (ground truth).

**Fig S8.**
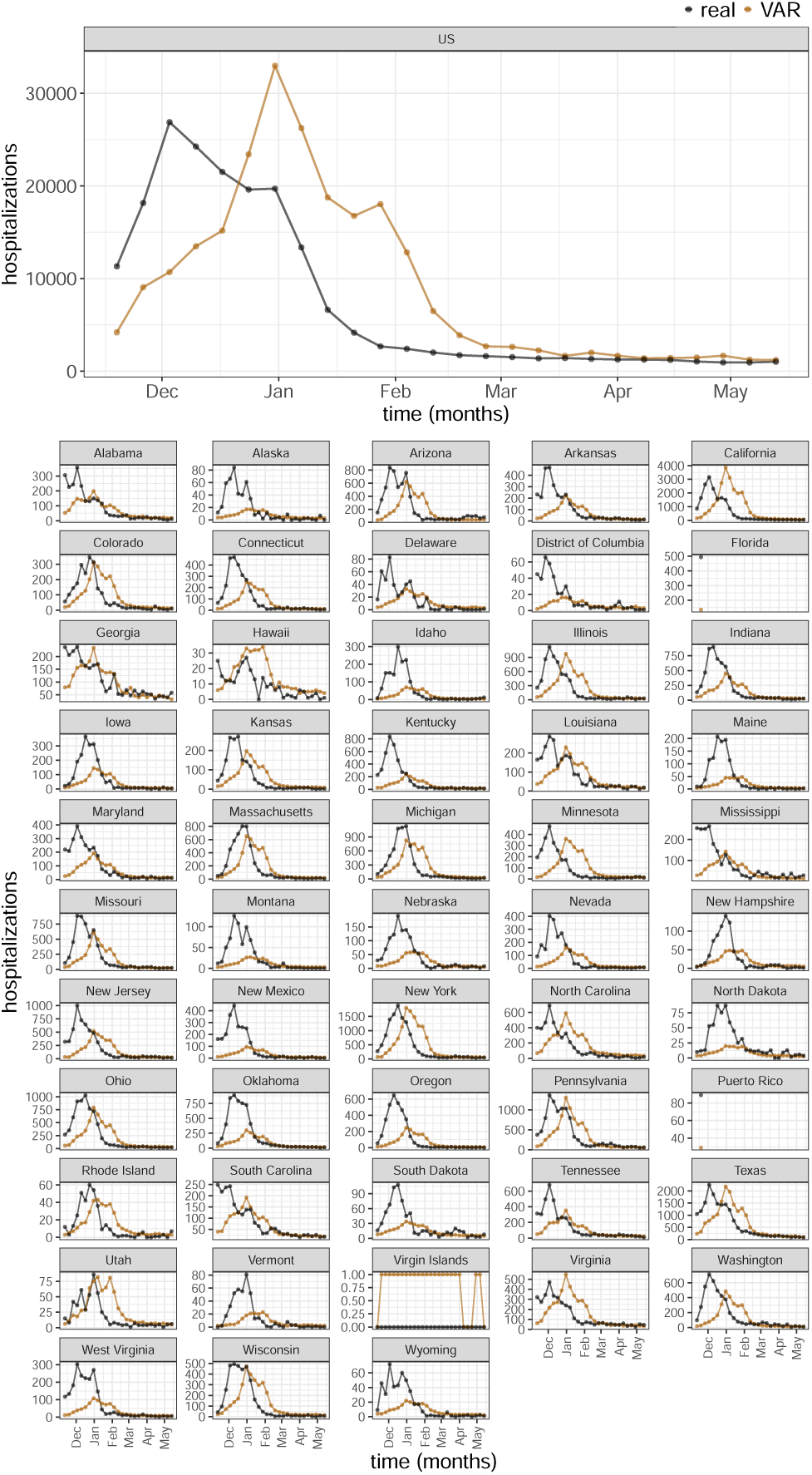
Comparison of VAR Model Forecasts with Ground Truth Data at Horizon 4 for the 2022-23 season. This figure illustrates the forecasting performance of of the regularized VAR model for the first prediction horizon (4 weeks ahead) relative to the actual observed hospitalizations (ground truth).

**Fig S9.**
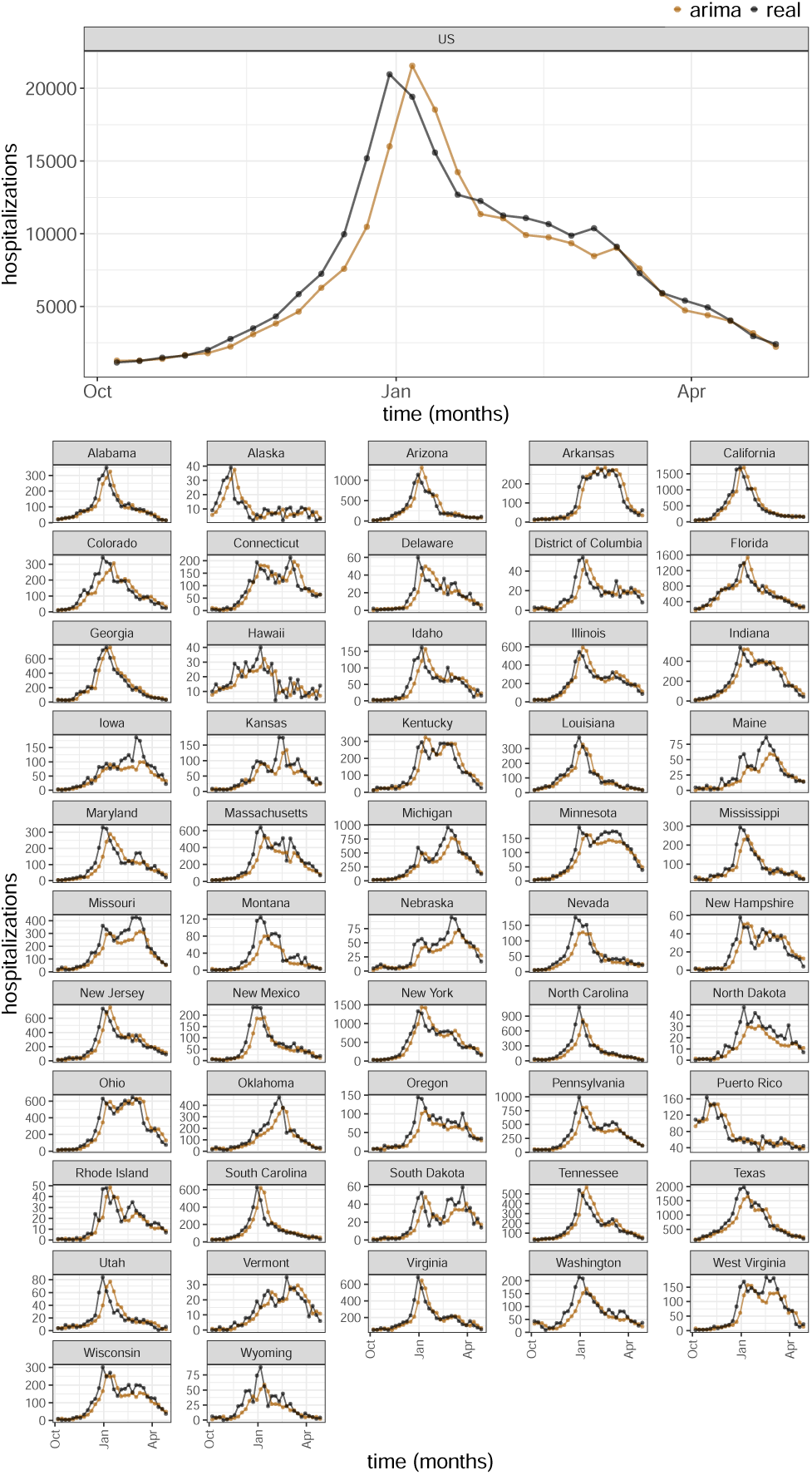
Comparison of ARIMA Model Forecasts with Ground Truth Data at Horizon 1 for the 2023-24 season. This figure illustrates the forecasting performance of the ARIMA model for the first prediction horizon (1 week ahead) relative to the actual observed hospitalizations (ground truth).

**Fig S10.**
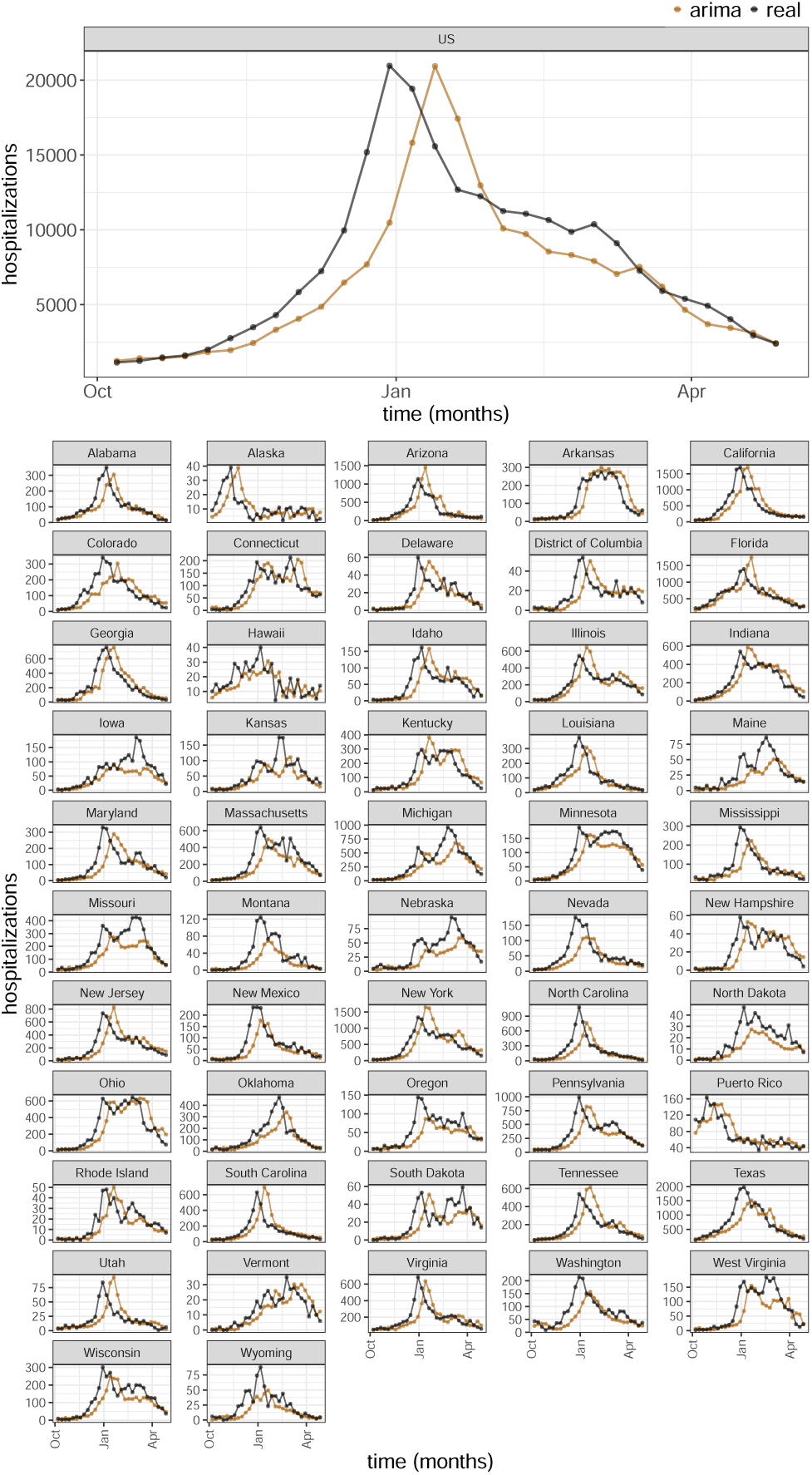
Comparison of ARIMA Model Forecasts with Ground Truth Data at Horizon 2 for the 2023-24 season. This figure illustrates the forecasting performance of the ARIMA model for the first prediction horizon (2 weeks ahead) relative to the actual observed hospitalizations (ground truth).

**Fig S11.**
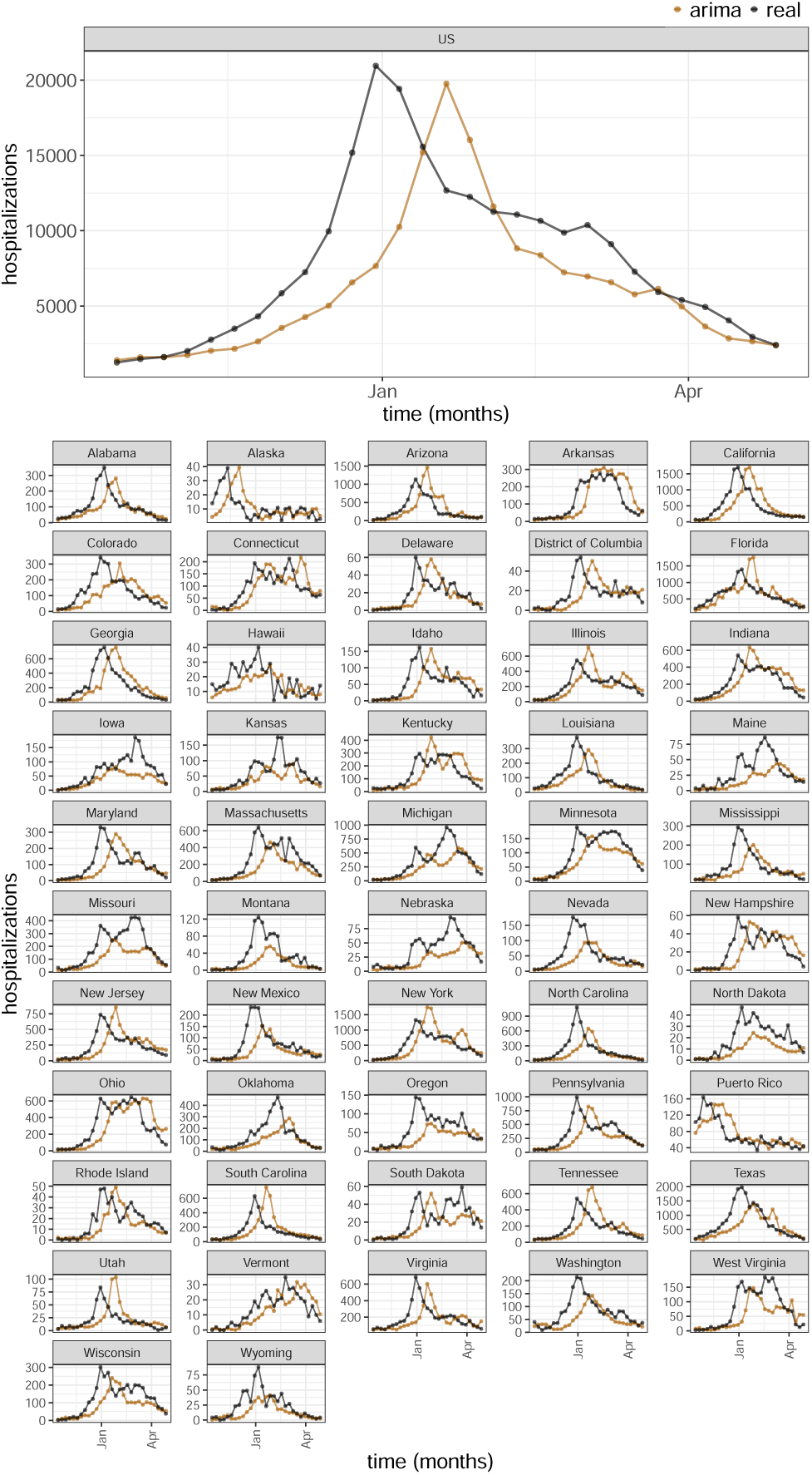
Comparison of ARIMA Model Forecasts with Ground Truth Data at Horizon 3 for the 2023-24 season. This figure illustrates the forecasting performance of the ARIMA model for the first prediction horizon (3 weeks ahead) relative to the actual observed hospitalizations (ground truth).

**Fig S12.**
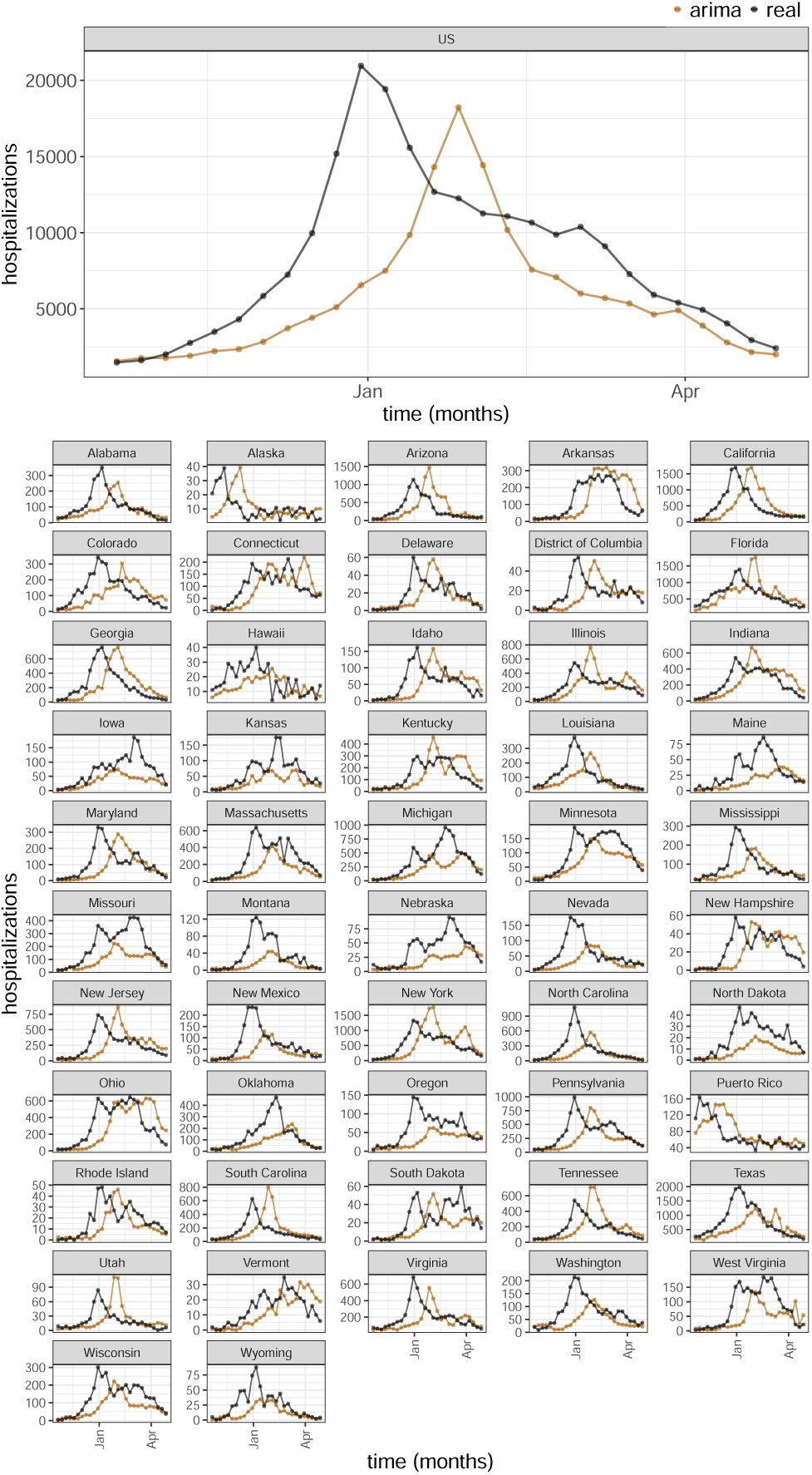
Comparison of ARIMA Model Forecasts with Ground Truth Data at Horizon 4 for the 2023-24 season. This figure illustrates the forecasting performance of the ARIMA model for the first prediction horizon (4 weeks ahead) relative to the actual observed hospitalizations (ground truth).

**Fig S13.**
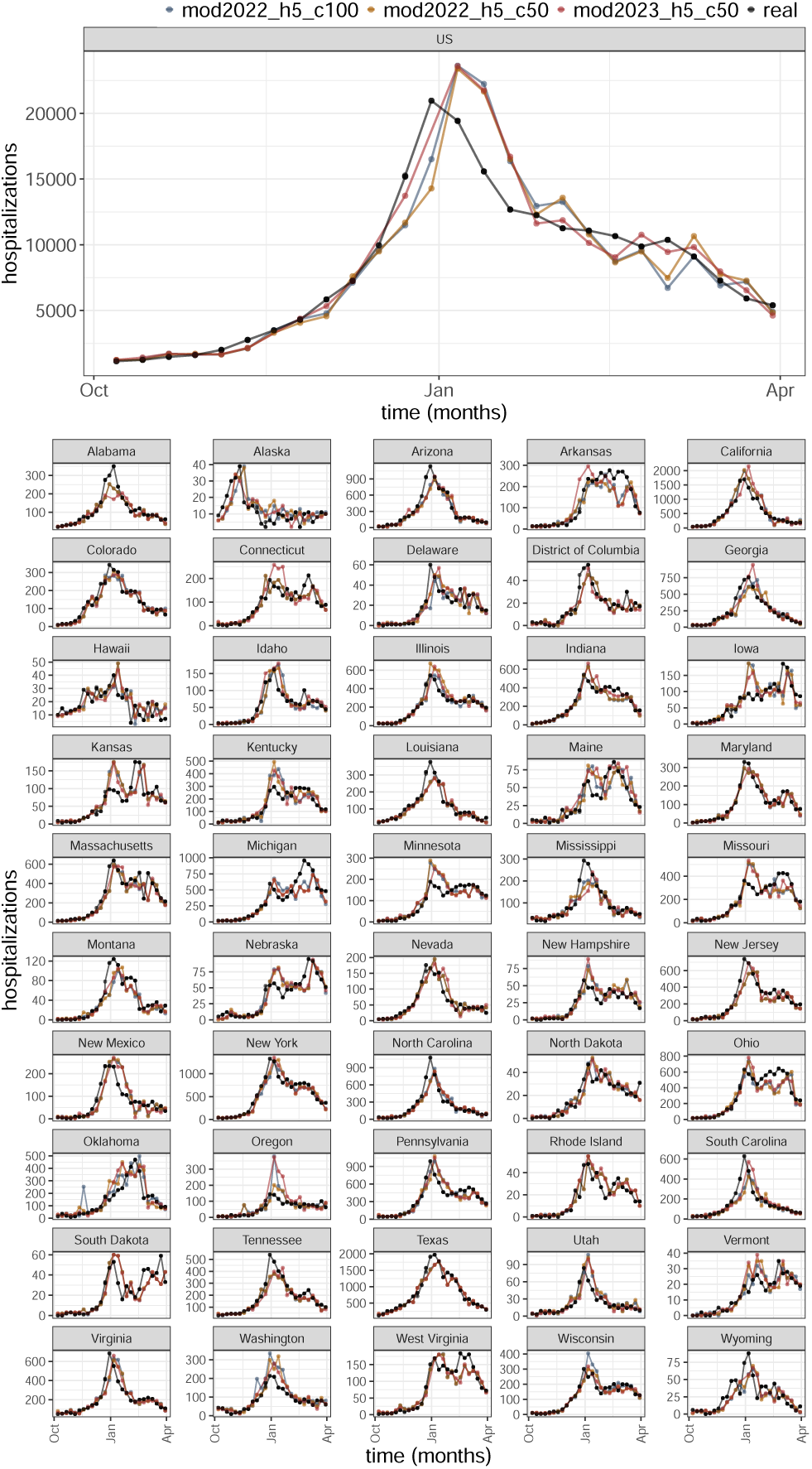
Comparison of LightGBM Model Forecasts with Ground Truth Data at Horizon 1 for the 2023-24 season. This figure illustrates the forecasting performance of each LightGBM model for the first prediction horizon (1 week ahead) relative to the actual observed hospitalizations (ground truth). The analysis includes models initiated with different random seeds and trained on distinct datasets up to June 2022 and June 2023, denoted as mod2022 h5 c100, mod2022 h5_c50, and mod2023_h5_c50, respectively.

**Fig S14.**
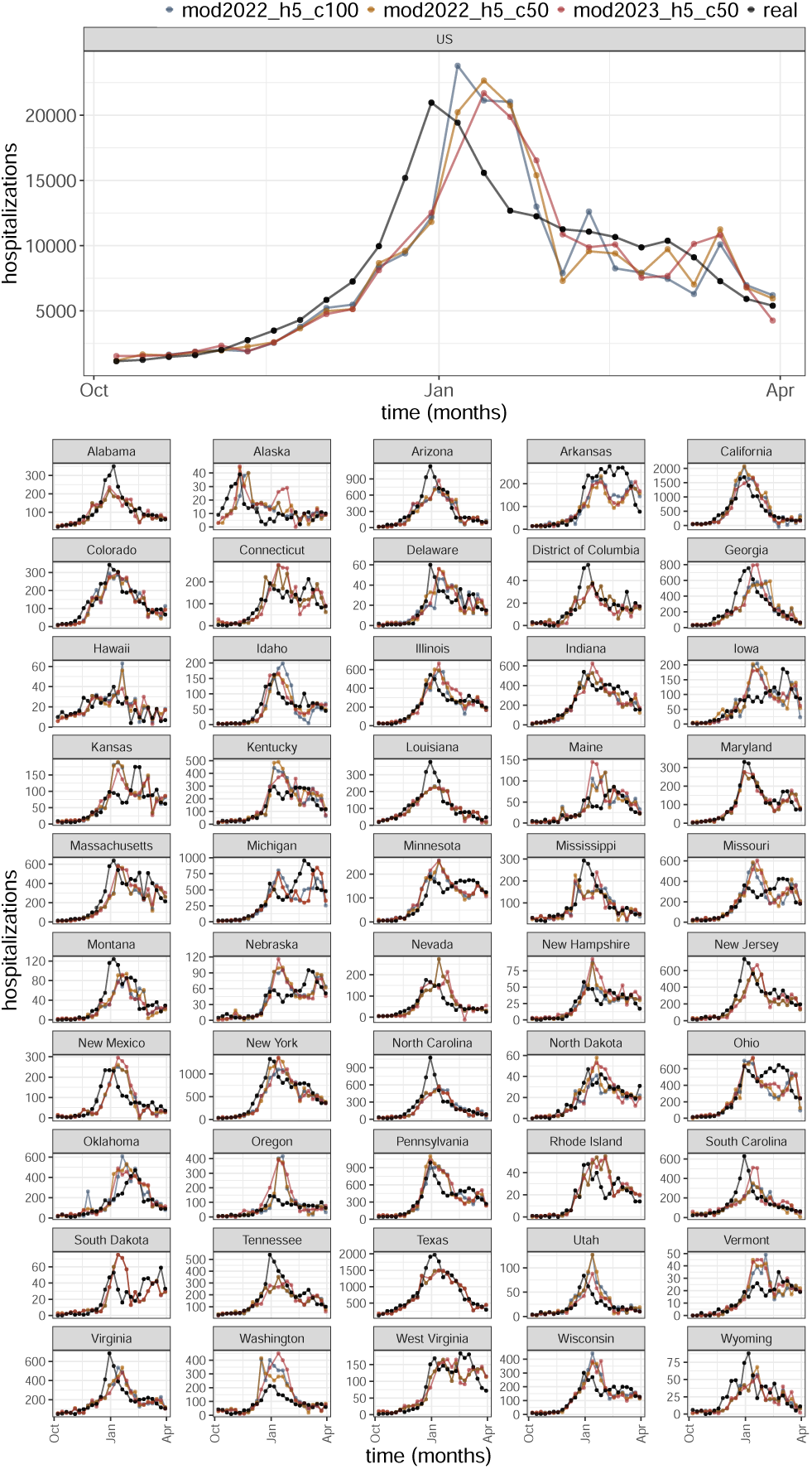
Comparison of LightGBM Model Forecasts with Ground Truth Data at Horizon 2 for the 2023-24 season. This figure illustrates the forecasting performance of each LightGBM model for the second prediction horizon (2 weeks ahead) relative to the actual observed hospitalizations (ground truth). The analysis includes models initiated with different random seeds and trained on distinct datasets up to June 2022 and June 2023, denoted as mod2022 h5 c100, mod2022 h5_c50, and mod2023_h5_c50, respectively.

**Fig S15.**
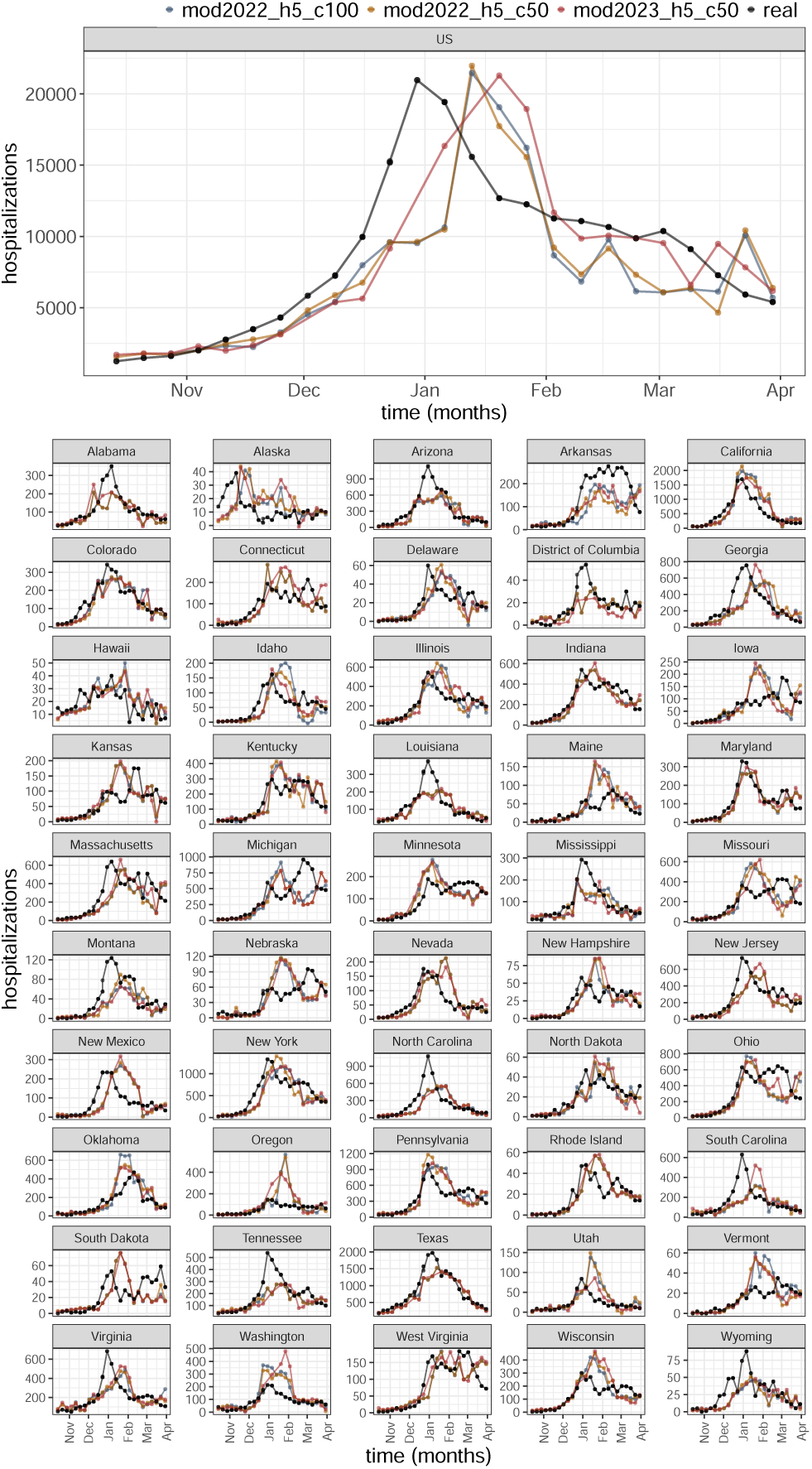
Comparison of LightGBM Model Forecasts with Ground Truth Data at Horizon 3 for the 2023-24 season. This figure illustrates the forecasting performance of each LightGBM model for the third prediction horizon (3 weeks ahead) relative to the actual observed hospitalizations (ground truth). The analysis includes models initiated with different random seeds and trained on distinct datasets up to June 2022 and June 2023, denoted as mod2022 h5 c100, mod2022 h5_c50, and mod2023_h5_c50, respectively.

**Fig S16.**
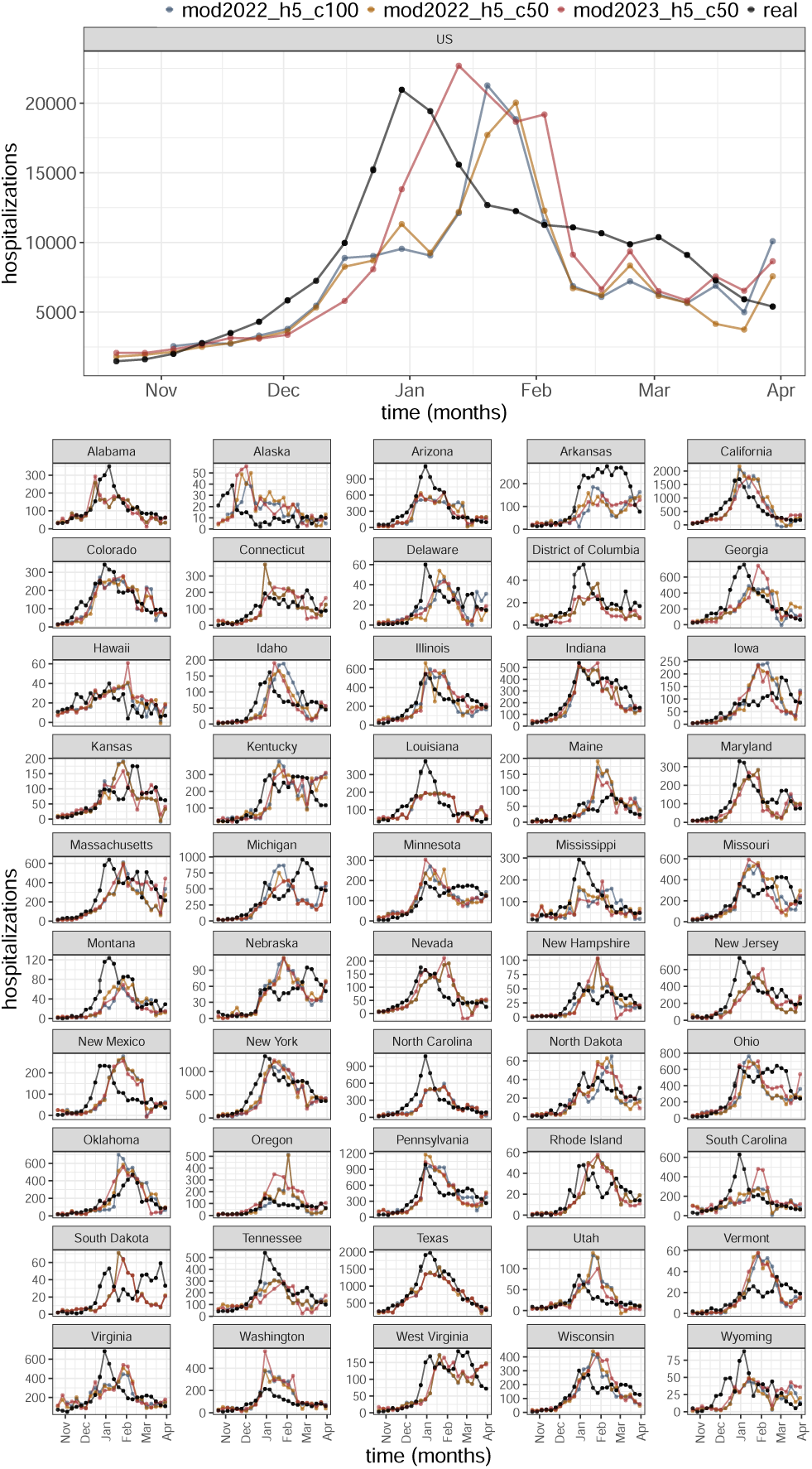
Comparison of LightGBM Model Forecasts with Ground Truth Data at Horizon 4 for the 2023-24 season. This figure illustrates the forecasting performance of each LightGBM model for the forth prediction horizon (4 weeks ahead) relative to the actual observed hospitalizations (ground truth). The analysis includes models initiated with different random seeds and trained on distinct datasets up to June 2022 and June 2023, denoted as mod2022 h5 c100, mod2022 h5_c50, and mod2023_h5_c50, respectively.

**Fig S17.**
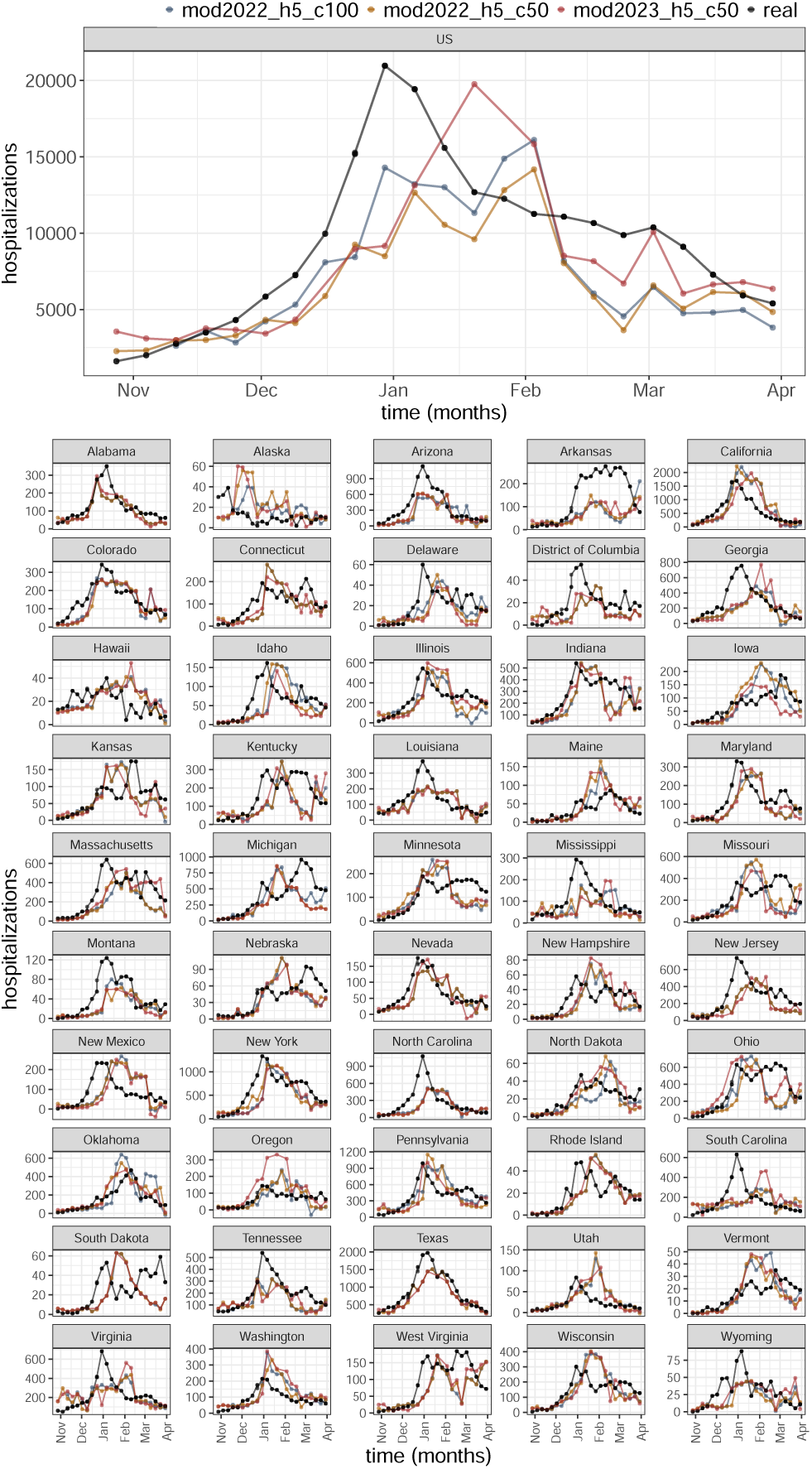
Comparison of LightGBM Model Forecasts with Ground Truth Data at Horizon 5 for the 2023-24 season. This figure illustrates the forecasting performance of each LightGBM model for the fifth prediction horizon (5 weeks ahead) relative to the actual observed hospitalizations (ground truth). The analysis includes models initiated with different random seeds and trained on distinct datasets up to June 2022 and June 2023, denoted as mod2022 h5 c100, mod2022 h5_c50, and mod2023_h5_c50, respectively.

## References

1. Centers for Disease Control and Prevention (CDC). Epidemiology and prevention of vaccine-preventable diseases. Hall E, Wodi AP, Hamborsky J, Morelli V, Schillie S, editors. Public Health Foundation; 2021.

2. Centers for Disease Control and Prevention (CDC). Influenza (Flu) Burden: Estimates of the Burden of Influenza from the Centers for Disease Control and Prevention;. https://www.cdc.gov/flu/about/burden/index.html, Accessed on 2024-04-10.

3. Gantenberg JR, McConeghy KW, Howe CJ, Steingrimsson J, Van Aalst R, Chit A, et al. Predicting Seasonal Influenza Hospitalizations Using an Ensemble Super Learner: A Simulation Study. American Journal of Epidemiology. 2023;192(10):1688–1700.

4. Yang S, Santillana M, Kou SC. Accurate estimation of influenza epidemics using Google search data via ARGO. Proceedings of the National Academy of Sciences. 2015;112(47):14473–14478.

5. Baltrusaitis K, Vespignani A, Rosenfeld R, Gray J, Raymond D, Santillana M, et al. Differences in regional patterns of influenza activity across surveillance systems in the United States: comparative evaluation. JMIR public health and surveillance. 2019;5(4):e13403.

6. Reich NG, Brooks LC, Fox SJ, Kandula S, McGowan CJ, Moore E, et al. A collaborative multiyear, multimodel assessment of seasonal influenza forecasting in the United States. Proceedings of the National Academy of Sciences. 2019;116(8):3146–3154.

7. Lu FS, Hattab MW, Clemente CL, Biggerstaff M, Santillana M. Improved state-level influenza nowcasting in the United States leveraging Internet-based data and network approaches. Nature communications. 2019;10(1):147.

8. Kandula S, Pei S, Shaman J. Improved forecasts of influenza-associated hospitalization rates with Google Search Trends. Journal of the Royal Society Interface. 2019;16(155):20190080.

9. Rangarajan P, Mody SK, Marathe M. Forecasting dengue and influenza incidences using a sparse representation of Google trends, electronic health records, and time series data. PLoS computational biology. 2019;15(11):e1007518.

10. Yang S, Ning S, Kou S. Use internet search data to accurately track state level influenza epidemics. Scientific reports. 2021;11(1):4023.

11. Ertem Z, Raymond D, Meyers LA. Optimal multi-source forecasting of seasonal influenza. PLoS computational biology. 2018;14(9):e1006236.

12. Ray EL, Reich NG. Prediction of infectious disease epidemics via weighted density ensembles. PLoS computational biology. 2018;14(2):e1005910.

13. Brooks LC, Farrow DC, Hyun S, Tibshirani RJ, Rosenfeld R. Nonmechanistic forecasts of seasonal influenza with iterative one-week-ahead distributions. PLoS computational biology. 2018;14(6):e1006134.

14. Ray EL, Sakrejda K, Lauer SA, Johansson MA, Reich NG. Infectious disease prediction with kernel conditional density estimation. Statistics in medicine. 2017;36(30):4908–4929.

15. Venkatramanan S, Sadilek A, Fadikar A, Barrett CL, Biggerstaff M, Chen J, et al. Forecasting influenza activity using machine-learned mobility map. Nature communications. 2021;12(1):726.

16. Ben-Nun M, Riley P, Turtle J, Bacon DP, Riley S. Forecasting national and regional influenza-like illness for the USA. PLoS computational biology. 2019;15(5):e1007013.

17. Osthus D, Gattiker J, Priedhorsky R, Del Valle SY. Dynamic Bayesian influenza forecasting in the United States with hierarchical discrepancy (with discussion). 2019;.

18. Aiken EL, Nguyen AT, Viboud C, Santillana M. Toward the use of neural networks for influenza prediction at multiple spatial resolutions. Science Advances. 2021;7(25):eabb1237.

19. Wu N, Green B, Ben X, O’Banion S. Deep transformer models for time series forecasting: The influenza prevalence case. arXiv preprint arXiv:200108317. 2020;.

20. Biggerstaff M, Alper D, Dredze M, Fox S, Fung ICH, Hickmann KS, et al. Results from the centers for disease control and prevention’s predict the 2013–2014 Influenza Season Challenge. BMC infectious diseases. 2016;16:1–10.

21. Biggerstaff M, Johansson M, Alper D, Brooks LC, Chakraborty P, Farrow DC, et al. Results from the second year of a collaborative effort to forecast influenza seasons in the United States. Epidemics. 2018;24:26–33.

22. McGowan CJ, Biggerstaff M, Johansson M, Apfeldorf KM, Ben-Nun M, Brooks L, et al. Collaborative efforts to forecast seasonal influenza in the United States, 2015–2016. Scientific reports. 2019;9(1):683.

23. Mathis SM, Webber AE, León TM, Murray EL, Sun M, White LA, et al. Evaluation of FluSight influenza forecasting in the 2021–22 and 2022–23 seasons with a new target laboratory-confirmed influenza hospitalizations. medRxiv. 2023;.

24. Ke G, Meng Q, Finley T, Wang T, Chen W, Ma W, et al. Lightgbm: A highly efficient gradient boosting decision tree. Advances in neural information processing systems. 2017;30.

25. Chaves SS, Lynfield R, Lindegren ML, Bresee J, Finelli L. The US influenza hospitalization surveillance network. Emerging infectious diseases. 2015;21(9):1543.

26. Centers for Disease Control and Prevention (CDC). Weekly U.S. Influenza Surveillance Report;. https://www.cdc.gov/flu/weekly/overview.htm, Accessed on 2024-04-10.

27. Peterson RA. Finding Optimal Normalizing Transformations via bestNormalize. The R Journal. 2021;13:294–313. doi:10.32614/RJ-2021-041.

28. Wickham H. ggplot2: Elegant Graphics for Data Analysis. Springer-Verlag New York; 2016. Available from: https://ggplot2.tidyverse.org.

29. Pedregosa F, Varoquaux G, Gramfort A, Michel V, Thirion B, Grisel O, et al. Scikit-learn: Machine Learning in Python. Journal of Machine Learning Research. 2011;12:2825–2830.

30. Löning M, Bagnall A, Ganesh S, Kazakov V, Lines J, Király FJ. sktime: A unified interface for machine learning with time series. arXiv preprint arXiv:190907872. 2019;.

31. Herzen J, Lässig F, Piazzetta SG, Neuer T, Tafti L, Raille G, et al. Darts: User-friendly modern machine learning for time series. Journal of Machine Learning Research. 2022;23(124):1–6.

